# Assessing the contribution of rare variants to congenital heart disease through a large-scale case-control exome study

**DOI:** 10.1101/2023.12.23.23300495

**Authors:** Enrique Audain, Anna Wilsdon, Gregor Dombrowsky, Alejandro Sifrim, Jeroen Breckpot, Yasset Perez-Riverol, Siobhan Loughna, Allan Daly, Pavlos Antoniou, Philipp Hofmann, Amilcar Perez-Riverol, Anne-Karin Kahlert, Ulrike Bauer, Thomas Pickardt, Sabine Klaassen, Felix Berger, Ingo Daehnert, Sven Dittrich, Brigitte Stiller, Hashim Abdul-Khaliq, Frances Bu’lock, Anselm Uebing, Hans-Heiner Kramer, Vivek Iyer, Lars Allan Larsen, J David Brook, Marc-Phillip Hitz

**Author notes:** Joint corresponding authors. These authors contributed equally to this work (Enrique Audain, Anna Wilsdon).

## Abstract

Several studies have demonstrated the value of large-scale human exome and genome data analysis, to maximise gene discovery in rare diseases. Using this approach, we have analysed the exomes of 4,747 cases and 52,881 controls, to identify single genes and digenic interactions which confer a substantial risk of congenital heart disease (CHD). We identified both rare loss-of-function and missense coding variants in ten genes which reached genome-wide significance (Bonferroni adjusted *P* < 0.05) and an additional four genes with a significant association at a false discovery rate (*FDR)* threshold of 5%. We highlight distinct genetic contributions to syndromic and non-syndromic CHD at both single gene and digenic level, by independently analysing probands from these two groups. In addition, by integrative analysis of exome data with single-cell transcriptomics data from human embryonic hearts, we identified cardiac-specific cells as well as putative biological processes underlying the pathogenesis of CHD. In summary, our findings strengthen the association of known CHD genes, and have identified additional novel disease genes and digenic interactions contributing to the aetiology of CHD.

## INTRODUCTION

Congenital Heart Disease (CHD) is a global health challenge, affecting ∼1-2% of live births worldwide^1^. However, despite advances in our understanding of the underlying disease aetiology in recent years, a significant proportion of CHD cases remains unexplained, suggesting that genetic mechanisms and other risk factors remain poorly understood^2,3^. Recent advances in exome and genome sequencing technologies have opened up new avenues of study and have resulted in novel insights into the genetic and epigenetic mechanisms underlying rare diseases, such as CHD^4,5^.

Previous studies have defined the association of inherited and *de novo* variations as a cause of CHD^6,7^. In addition, these studies have highlighted the differences between the genetic architecture of syndromic (with extracardiac malformations and/or neurodevelopmental delay) and non-syndromic (isolated) CHD^6,7^. Continuing collaboration between the scientific community and healthcare teams has driven efforts to integrate and analyse larger cohorts of patients, and has demonstrated the potential of this approach to uncover novel variants and genes associated with CHD^6–8^.

Here, we present a whole exome sequencing analysis of 4,747 CHD cases and 52,881 controls. This is one of the largest cohorts of non-syndromic CHD cases (n=2,929) studied so far, meaning that we are in an advantageous position to refine our understanding of the genetic mechanisms underlying non-syndromic CHD specifically. This is especially important given that the vast majority of individuals with CHD, have non-syndromic CHD.

We used the case-control cohort to investigate both single genes and digenic interactions contributing to CHD. We integrated data obtained in the case-control study with single-cell transcriptome data obtained from human embryonic hearts^9^. This complementary analysis identified biological processes enriched for genes differentially expressed in cardiac-specific cells, found also significant in our case-control analysis. Importantly, the data suggest a difference in cardiac developmental mechanisms between syndromic and non-syndromic CHD.

Taken together, we have identified ten genome-wide significant (*Bonferroni adjusted P < 0.05*) genes, and an additional four genes at *FDR* 5%, which are associated with CHD, as well as a larger contribution of digenic interactions to non-syndromic compared to syndromic CHD.

## RESULTS

### Cohort description and analysis workflow

We combined and analysed the exomes of 4,747 CHD cases (aCHD, refers to all CHD cases) and 52,881 controls. CHD cases were further classified into syndromic CHD (sCHD, individuals with extracardiac malformations or neurodevelopmental disability, n=1,818) and non-syndromic CHD (nsCHD, individuals with isolated CHD, n=2,929). All samples and genetic variants were subjected to a sequence of quality control steps to obtain a final cohort of unrelated and matched-ancestry individuals, as well as a set of high-confidence variants for downstream analysis (see **Methods, Supplemental Information 1**).

We evaluated the distribution of high-confidence loss-of-function (hcLOF) and missense constrained variants (missC) across a spectrum of LOF and missense constrained genes (**Methods**). In addition, we performed gene-based burden testing to identify genes conferring a high risk of CHD, as well as the expression pattern at single-cell resolution. Lastly, we evaluate the contribution of digenic interactors to syndromic and non-syndromic CHD. **Figure 1** simplifies the workflow followed in this study to discover novel associations with CHD.

**Figure 1.**
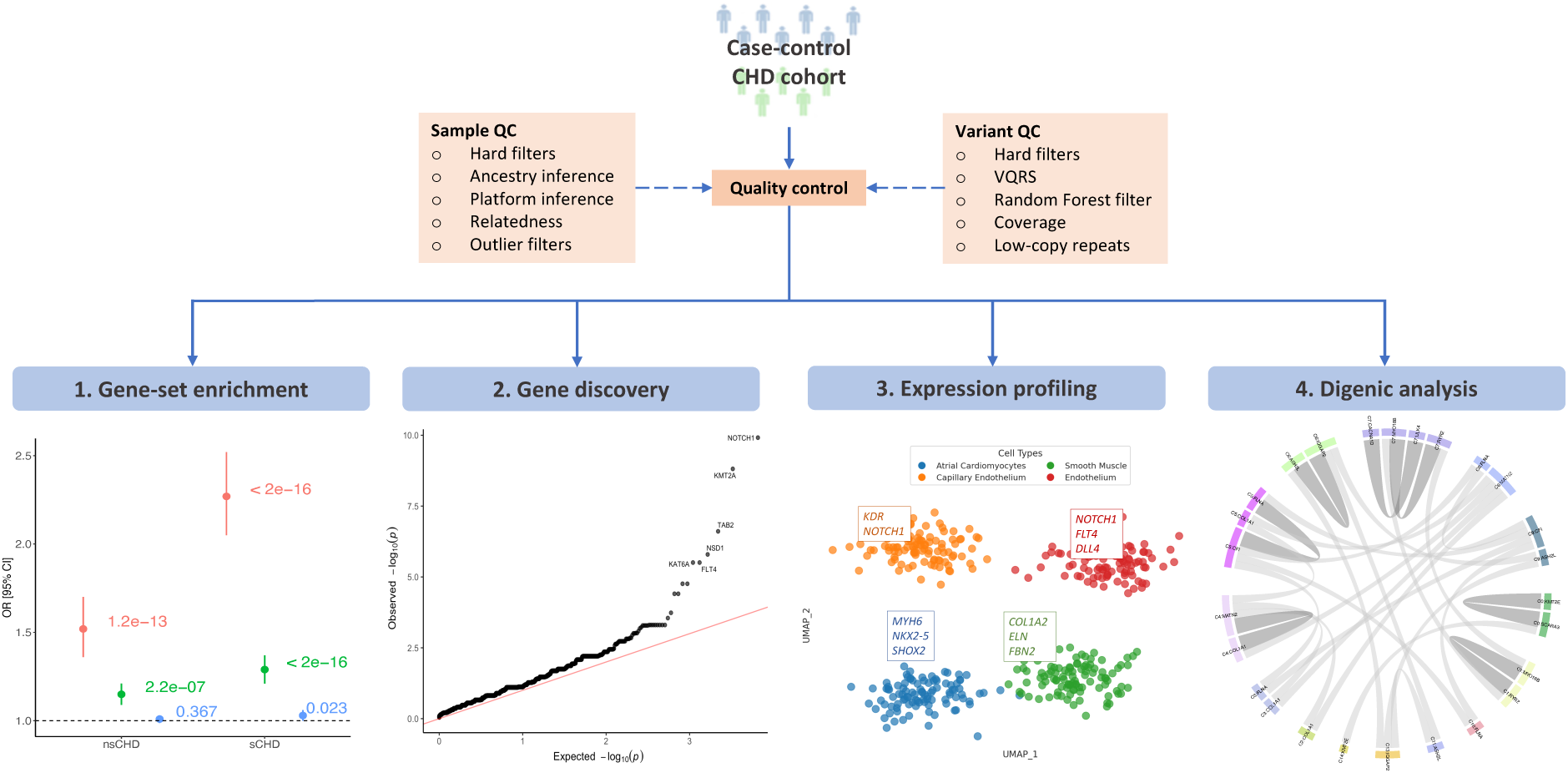
Analysis workflow for disease gene discovery. Quality control processes were conducted at the sample and variant levels. **(1)** Gene-set enrichment analysis was performed on the gene intolerance to missense and loss-of-function spectrum. **(2)** Gene-based case-control burden testing (Fisher’s Exact test) was performed for high-confidence loss-of-function (hcLOF) and missense constrained variants (missC) independently. The per gene minimal *p-value* (*P*) from both analyses was set as the study-wide *p-value*, corrected for multiple testing using the Bonferroni and B-H methods. **(3)** Expression profiling of significant CHD genes differentially expressed on cardiac specific cell clusters **(4)** Digenic analysis was conducted by comparing the rate of mutations observed on cases compared to controls. All analysis were stratified by syndromic status (aCHD, sCHD and nsCHD) vs control.

### Distinct pattern of loss-of-function constrained genes identified between sCHD and nsCHD

Previous studies have suggested a greater contribution of loss-of-function (LOF) variants to sCHD, compared to non-syndromic forms^6,7^. To determinate if this holds true in this present cohort, we evaluated the burden of rare variants in the sCHD and nsCHD cohort, compared with controls across the per gene spectrum of loss-of-function intolerance. Following the approach proposed by the gnomAD consortium^10^, we divided 19,923 protein-coding genes into ten bins (∼1,900 genes per bin) based on its observed/expected LOF ratio upper fraction (termed LOEUF) and applied a logistic regression model (see **Methods**) to each bin (i.e., gene-set). This allowed us to assess enrichment across three different functional categories of variants (hcLOF, missC and synonymous), stratified by CHD probands (aCHD, sCHD and nsCHD). The highest enrichment was observed in the most LOF constrained genes (bin 1) for hcLOF variants (**Figure 2**). These variants provided a major contribution to sCHD cases (*OR* = 2.27, *P* < 2 × 10^-16^), and much less so for nsCHD (*OR* = 1.52, *P* = 1.2 × 10^-13^). A moderate enrichment was observed for missC variations, suggesting that this class of variants could have a similar (although smaller) functional impact compared to hcLOF variants. Although reduced in magnitude, this same pattern was also observed in the set of genes in the second LOEUF constraint bin, whereas no enrichment was observed towards less LOEUF constrained bins (**Figure 2**). No enrichment of synonymous variants was observed across the bins, providing a negative control set (**Figure 2)**.

**Figure 2.**
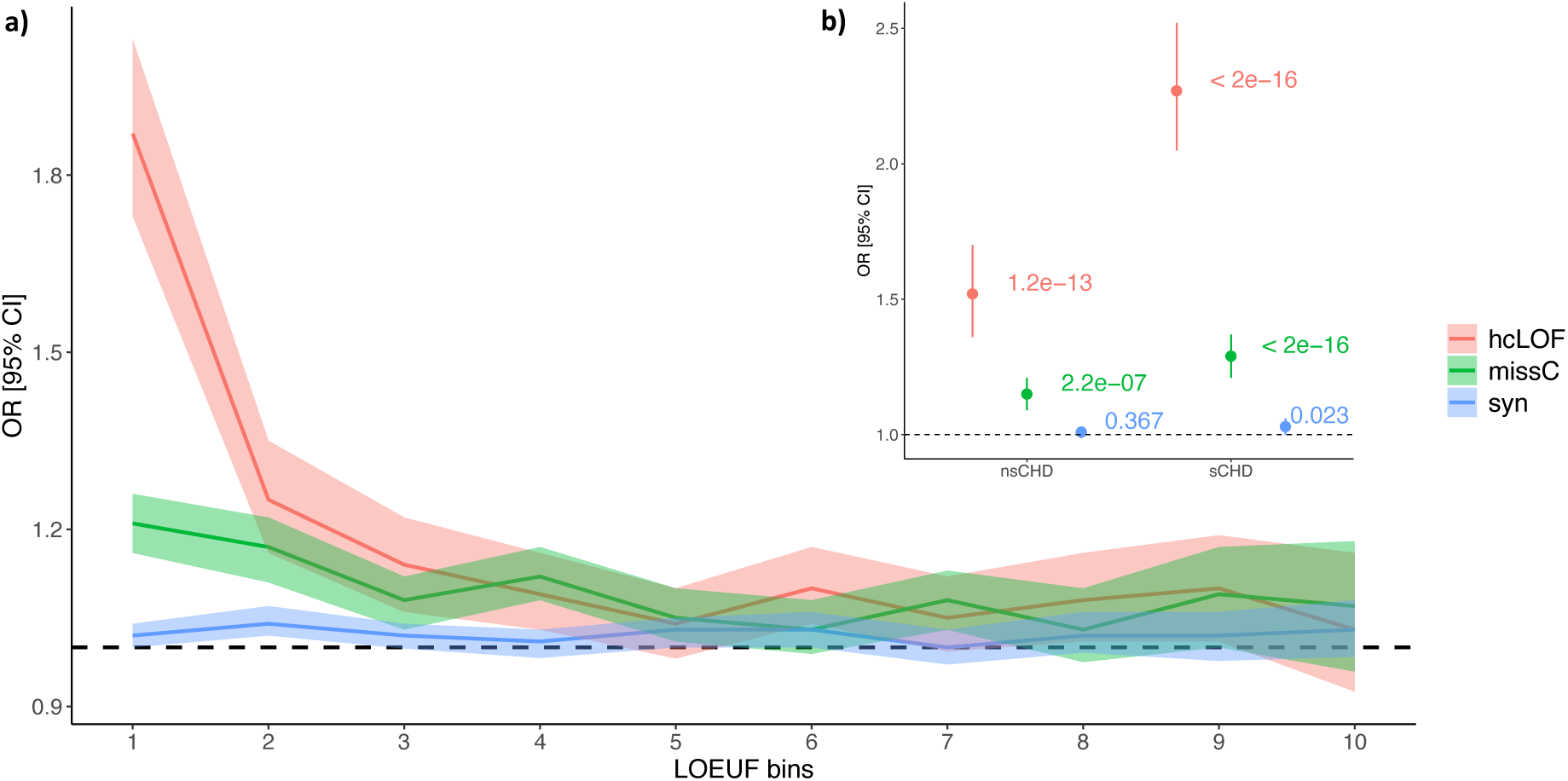
Enrichment analysis across the LOF constraint gene spectrum. Protein-coding genes were binned based on the LOEUF metric as proposed by gnomAD. Every bin contains ∼1,900 genes. Top bins (1, 2) contain genes with the highest intolerance to loss-of-function. **a)** Enrichment analysis comparing aCHD vs controls. **b)** Enrichment analysis stratified by syndromic status (sCHD and nsCHD) vs controls in the top constraint LOF bin (1). The x-axis indicates the constraint bins; the y-axis shows the Odd Ratios (OR) and the 95% confidence interval.

When the same analysis was performed across the missense constraint spectrum, assessed by the observed/expected missense ratio upper fraction gene-based metric (termed MOEUF), a similar pattern as described above (higher enrichment in the most missense-constrained genes) was observed (**Supplemental Figure 1**).

These results demonstrate a larger effect of hcLOF compared to missC variants across the LOEUF and MOEUF spectrum, with the major contribution observed in sCHD, compared with nsCHD. Nevertheless, the results suggest that both hcLOF and missC variants are important genetic components contributing to CHD development.

### Gene-based enrichment analysis

To identify genes that confer a significant risk of CHD, we performed a case-control burden analysis by combining rare variants (MAF < 0.001) at the gene level. It has been demonstrated that following a method of collapsing variants within specific genomic regions (e.g., genes), increases the power to discover new associations at low allele frequencies^11^. Following this principle, we conducted a Fisher’s exact test to identify genes with a significant burden of non-synonymous variants in CHD cases compared to controls, and evaluated them independently for sCHD and nsCHD.

As with earlier comparable case-control exome studies^12–14^, the burden test was performed separately for hcLOF (*P_lof_*) and missC (*P_miss_*), and the minimal *p-value* observed per gene between these two variant categories was selected as the study-wide *p-value* (*P*). hcLOF variants were defined using the LOFTEE tool^10^, whereas missense variants were defined based on different missense deleteriousness prediction scores (see **Methods, Supplemental Figure 2**). Ten genes were identified with significant *P*, after correcting for multiple testing using the Bonferroni method (**Table 1, Supplemental Table 1**). Eight genes were associated with sCHD (*KMT2A*, *SMAD4*, *PTPN11, TAB2, NSD1, BCOR, KAT6A, PBX1)* and two were identified through the nsCHD (*FLT4* and *NOTCH1*) analysis. In addition, four genes showed significant associations with CHD at *FDR* 5% (*CTCF, KAT6B, SHOX2, HCAR1*). The evaluation of the set of synonymous variants showed a similar distribution of expected vs observed p-values, suggesting no genomic inflation of the test statistic (**Supplemental Figure 3**).

**Table 1.** Top 21 genes in the case-control burden analysis using the Fisher Exact test stratified by syndromic status (sCHD and nsCHD). A total of 16,351 genes were tested per variant type (hcLOF and missC). Analysis: sCHD or nsCHD vs controls. Consequence: denotes the consequence group with the minimal p-value (*P*). sCHD: number of syndromic cases (heterozygous). nsCHD: number of non-syndromic cases (heterozygous). Controls: number of controls (heterozygous). *P*: the minimal p-value per gene between *P_lof_* and *P_miss_.* P adj (FDR): Adjusted minimal p-value (*P*) using the B-H method with n = 2*16,351. P adj (Bonferroni): Adjusted minimal p-value (*P*) using the Bonferroni method with n = 2*16,351. In bold are highlighted the ten genes with *Bonferroni adjusted P* < 0.05. **Supplemental Table 1** contains the results for all protein-coding genes tested.

| Genes | Analysis | Consequence | sCHD | nsCHD | Controls | $P$ | $P$ adj (FDR) | $P$ adj (Bonferroni) |
| --- | --- | --- | --- | --- | --- | --- | --- | --- |
| **KMT2A** | sCHD | hcLOF | 8 | 0 | 0 | 9.76E-13 | 3.19E-08 | 3.19E-08 |
| **SMAD4** | sCHD | missC | 11 | 3 | 16 | 2.47E-10 | 4.04E-06 | 8.09E-06 |
| **NOTCH1** | nsCHD | hcLOF | 2 | 7 | 0 | 8.48E-10 | 2.77E-05 | 2.77E-05 |
| **PTPN11** | sCHD | missC | 11 | 5 | 25 | 8.78E-09 | 9.57E-05 | 2.87E-04 |
| **TAB2** | sCHD | hcLOF | 5 | 1 | 0 | 3.13E-08 | 2.56E-04 | 1.02E-03 |
| **NSD1** | sCHD | hcLOF | 5 | 1 | 1 | 1.83E-07 | 1.20E-03 | 5.98E-03 |
| **BCOR** | sCHD | hcLOF | 4 | 0 | 0 | 9.93E-07 | 4.06E-03 | 3.25E-02 |
| **KAT6A** | sCHD | hcLOF | 4 | 1 | 0 | 9.93E-07 | 4.06E-03 | 3.25E-02 |
| **PBX1** | sCHD | missC | 6 | 3 | 6 | 7.73E-07 | 4.06E-03 | 2.53E-02 |
| **FLT4** | nsCHD | hcLOF | 0 | 5 | 0 | 3.32E-07 | 5.43E-03 | 1.09E-02 |
| **CTCF** | sCHD | missC | 4 | 1 | 1 | 4.84E-06 | 1.58E-02 | 1.58E-01 |
| **KAT6B** | sCHD | hcLOF | 4 | 1 | 1 | 4.84E-06 | 1.58E-02 | 1.58E-01 |
| **SHOX2** | nsCHD | missC | 1 | 10 | 21 | 1.81E-06 | 1.98E-02 | 5.93E-02 |
| **HCAR1** | nsCHD | missC | 2 | 9 | 18 | 4.40E-06 | 3.60E-02 | 1.44E-01 |
| **ADNP** | sCHD | hcLOF | 3 | 0 | 0 | 3.15E-05 | 6.44E-02 | 1.00E+00 |
| **CHD7** | sCHD | hcLOF | 3 | 0 | 0 | 3.15E-05 | 6.44E-02 | 1.00E+00 |
| **EP300** | sCHD | hcLOF | 3 | 1 | 0 | 3.15E-05 | 6.44E-02 | 1.00E+00 |
| **KMT2D** | sCHD | hcLOF | 3 | 0 | 0 | 3.15E-05 | 6.44E-02 | 1.00E+00 |
| **KRT25** | sCHD | missC | 8 | 3 | 31 | 2.51E-05 | 6.44E-02 | 8.19E-01 |
| **QRICH1** | sCHD | hcLOF | 3 | 0 | 0 | 3.15E-05 | 6.44E-02 | 1.00E+00 |
| **SLC38A9** | nsCHD | missC | 0 | 6 | 6 | 1.19E-05 | 7.78E-02 | 3.89E-01 |

Of the genes identified as significant in sCHD, *KMT2A* (AD Wiedemann-Steiner syndrome OMIM 159555) showed the highest enrichment (**Figure 3a**). *NOTCH1* (AD Adams-Oliver syndrome 5, Aortic valve disease 1 OMIM 190198) showed the highest number of variations in the nsCHD cohort (**Figure 3b**) and warranted further investigation (companion manuscript).

**Figure 3.**
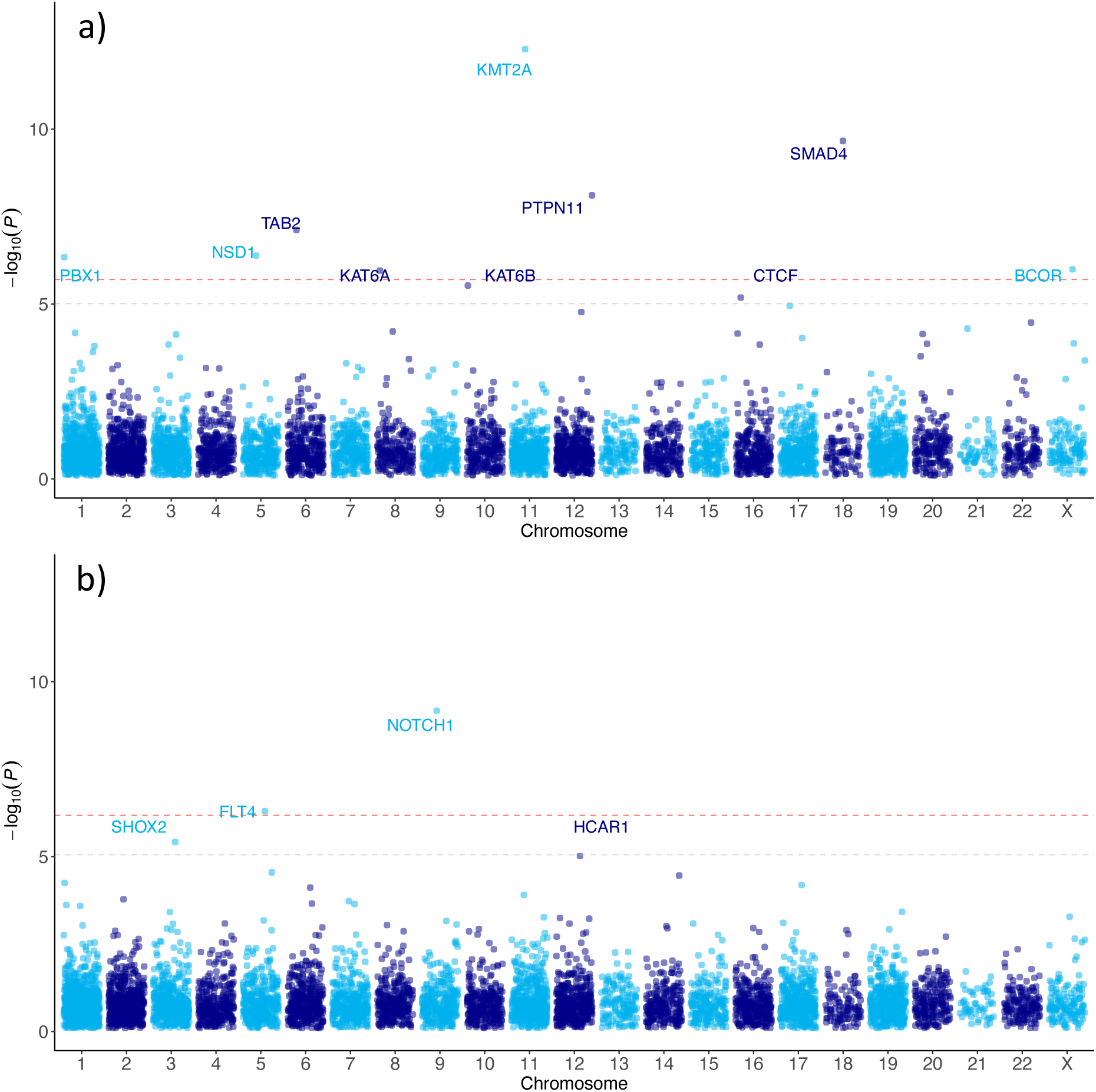
Log-transformed minimal p-value (*P*) per gene (y-axis) against its chromosomal location (x-axis). Red dashed line denotes the threshold for genes reaching exome-wide significance (Bonferroni adjusted *P* < 0.05); grey dashed line marks the threshold for genes reaching suggestive exome-wide significance (FDR 5%). a) Burden analysis of sCHD vs controls; b) burden analysis of nsCHD vs controls.

Other genes reaching a significant level of association included *NSD1* (AD Sotos Syndrome OMIM 606681), *TAB2* (AD Non-Syndromic CHD 2, OMIM 605101), *KAT6A* (AD Arboleda-Tham Syndrome OMIM 601408), *PTPN11* (AD Noonan Syndrome OMIM 176876), *SMAD4* (AD Mhyre Syndrome OMIM 600993), *FLT4* (AD Congenital heart defects, multiple types, 7 OMIM 136352), and the X-linked gene *BCOR* (XLD Syndromic Micropthalmia OMIM 300485). They have all been previously described in the context of CHD, and our results corroborate these findings.

The association of *PBX1* (AD Congenital anomalies of kidney and urinary tract syndrome with or without hearing loss, abnormal ears, or developmental delay OMIM 176310), *CTCF* (AD Intellectual developmental disorder, autosomal dominant 21 OMIM 604167) and *KAT6B* (AD Genitopatellar syndrome and SBBYSS syndrome OMIM 605880) with CHD (**Table 1**) have been previously reported in isolated cases or small patient cohorts, and our results add further evidence for an association with CHD.

*HCAR1* (OMIM 606923) and *SHOX2* (OMIM 602504) have not previously been associated with CHD at a genome-wide level. However, both genes were significantly associated with nsCHD at *FDR* 5% (**Figure 3b**).

### Differentially expressed genes in cardiac-specific cells show a distinct enrichment pattern in syndromic and non-syndromic CHD

Previous studies have revealed significant levels of expression in the heart of genes associated with CHD^7,8^. By using publicly accessible bulk RNAseq data^15^ (**Methods**), we consistently showed that genes with significant level of association in our case-control analysis also showed high expression in cardiac tissues (**Supplemental Figure 4**). Moreover, syndromic CHD genes showed a systematic elevated expression in other tissues (e.g., brain and kidney), compared to non-syndromic CHD (**Supplemental Figure 5**). The difference in expression patterns between these two groups was negligible in the heart, though (*P* > 0.05, Wilcoxon test; **Supplemental Figure 5**). Despite its relevance, bulk RNAseq data analysis does not stretch as far as the delineation of expression patterns at the cellular level.

To accomplish this, we assessed the mutational burden of rare non-synonymous variants (hcLOF and missC) within differentially expressed genes (DEGs) in cardiac-specific cells. We meta-analysed the exome data with a publicly available human heart transcriptomic dataset generated from early developmental stages of the human heart (6.5 and 7 weeks post-conception)^9^. Using the logistic regression framework mentioned above, we performed gene-set enrichment analysis on DEGs defined on 15 distinct cardiac cell clusters (C0-C14) reported by Asp *et al*^9^. Both hcLOF and missC mutations were evaluated independently and the analysis was stratified further by proband CHD status versus controls (aCHD, sCHD and nsCHD, **Figure 4**).

**Figure 4.**
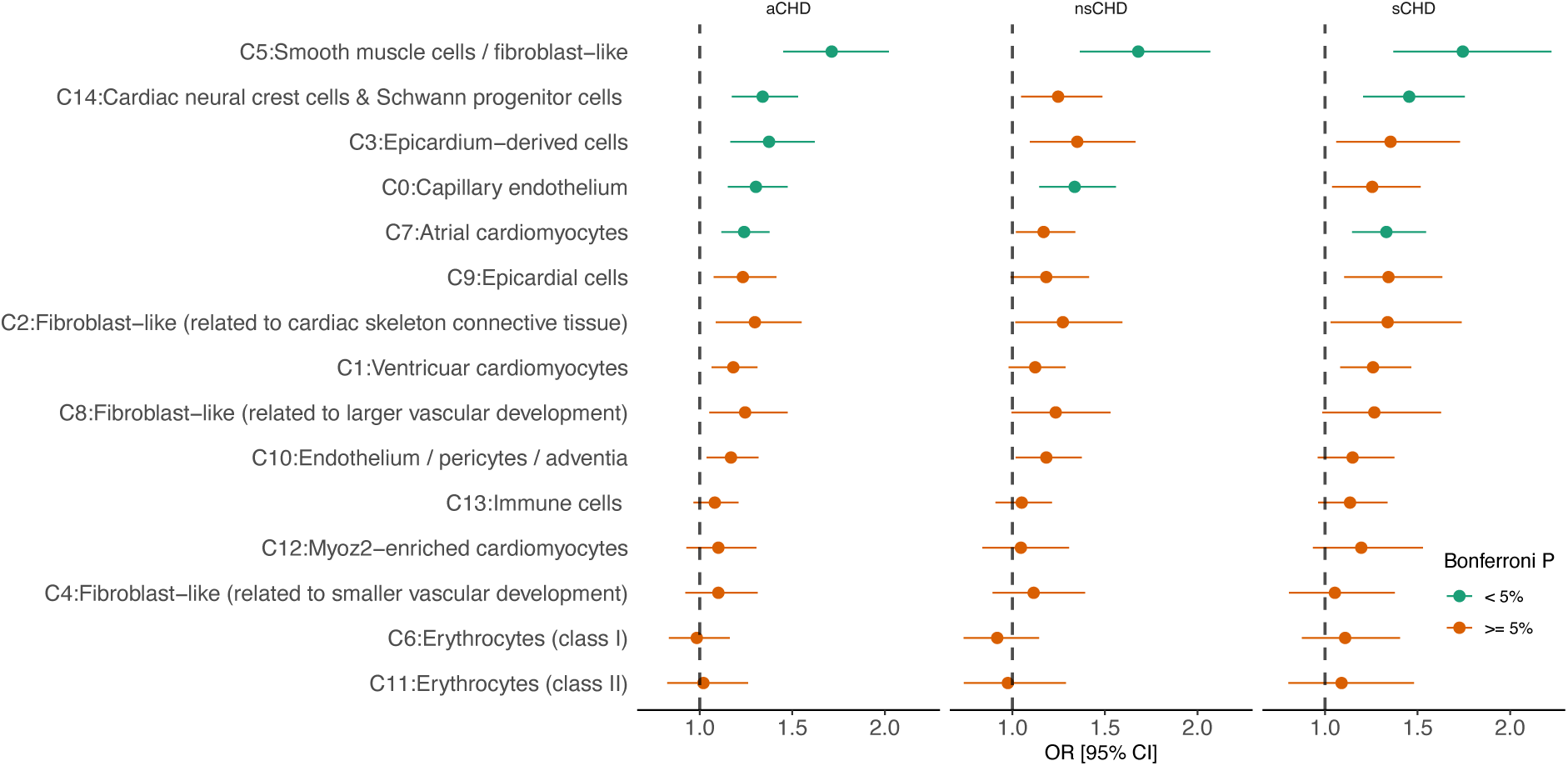
Logistic regression-based enrichment analysis of differentially expressed genes (DEGs) in cardiac-specific cell clusters for high-confidence loss-of-function variants (hcLOF). The analysis was stratified by syndromic status (aCHD, sCHD and nsCHD). The x-axis denotes the Odd Ratio (OR) and the 95% confidence interval. *P-values* were adjusted using the Bonferroni method (0.05 / 45 tests) to assess significant enrichment. Cardiac cell clusters C0, C3, C5, C7 and C14, show significant enrichment when analysing aCHD vs controls. The enrichment observed in clusters C7 and C14 showed a major contribution of sCHD. In comparison, cluster C0 provided the major contribution to nsCHD.

Five cardiac-specific cell clusters were found significantly enriched (Bonferroni adjusted *P* < 0.05) for hcLOF variations when analysing aCHD probands vs controls (**Figure 4**): Smooth muscle cells (C5), Cardiac neural crest cells (C14), Epicardium-derived cells (C3), Capillary endothelium (C0) and Atrial cardiomyocytes (C7). Enrichment of hcLOF variants for DEGs in Smooth muscle cells (C5) showed a significant contribution to both sCHD and nsCHD. Cardiac neural crest cells (C14) and Atrial cardiomyocytes (C7) contributed to sCHD in the main, whereas the cluster of Capillary endothelium cells was significantly enriched in nsCHD versus controls (**Figure 4**).

A similar enrichment pattern was observed when analysing the set of missC variants (**Supplemental Figure 6**). In addition to the Capillary endothelium (C0), Smooth muscle cells (C5), Atrial cardiomyocytes (C7) and Cardiac neural crest cells (C14) clusters; which were also found significantly enriched for hcLOF variants; two other cardiac-specific cell clusters showed a significant burden of missC variants in CHD cases (aCHD) compared to controls: Endothelium/pericytes cells (C10) and Fibroblast cells (C2).

The synonymous variants set was used as a negative control and did not identify enrichment in any clusters evaluated (Bonferroni adjusted *P* > 0.05, **Supplemental Figure 7**).

Together, these results provide valuable evidence regarding the possible mechanisms involved in the pathogenesis of CHD.

### Gene Ontology (GO) enrichment analysis

To provide additional supporting evidence for our previous findings, we performed Gene Ontology (GO) enrichment analysis to identify relationships between the enriched DEGs in cardiac-specific cell clusters to biological processes. We analysed the set of DEGs with an unadjusted *P* < 0.01 (Fisher Exact test) identified in the case-control burden analysis within the cell clusters showing enrichment in either the aCHD, sCHD or nsCHD analysis (**Supplemental Figure 8**).

Among the DEGs in cardiac-specific cells evaluated with the Enrichr tool^16^ (see **Methods**), four clusters showed at least one GO term with *FDR* < 1%. The data suggested that cell cluster C7 (Atrial cardiomyocytes, **Supplemental Figure 8a**) was mainly associated with biological processes involved in developing cardiac muscle tissue, and the observed signal was driven by *NKX2-5, MYH6, MYOCD, PKP2, BMP7, ANKRD1* and *ACTC1.* DEGs in C0 (Capillary endothelium, **Supplemental Figure 8b**) showed enrichment for vasculogenesis, with contribution from *KDR*, *NOTCH1* and *RASIP1*. C5 (Smooth muscle cells, **Supplemental Figure 8c**) was associated with extracellular matrix organisation processes, with a noteworthy contribution of genes that contain a collagen-like domain (e.g., *COL14A1* and *COL1A2*), as well as *ELN* and *FBN2.* DEGs in C10 (Endothelium and pericyte cells, **Supplemental Figure 8d**), demonstrated the higher enrichment of missC variants (**Supplemental Figure 6**), and enrichment of biological process involved in the cellular response to vascular endothelial growth factor stimulus and the regulation of cell migration as part of sprouting angiogenesis. *DLL4, FLT4, KDR, MEOX2* and *NOTCH1* all contributed to this cluster.

### Contribution of digenic interactions to syndromic and non-syndromic CHD

Next, we studied the contribution of digenic interactions to CHD using the RareComb^17^ framework (**see Methods**). Our analysis revealed a total of 2,083 digenic pairs significantly enriched for hcLOF and/or missC variants in CHD cases (aCHD) compared to controls at *FDR* 1% (**Figure 5a, Supplemental Table 2**). The data suggested that a significantly higher proportion of digenic interactors contributed to non-syndromic (n=810) forms of CHD (*P* = 6.7 × 10^-3^, proportion Z-test, **Figure 5a**) compared to syndromic forms (n=433).

**Figure 5.**
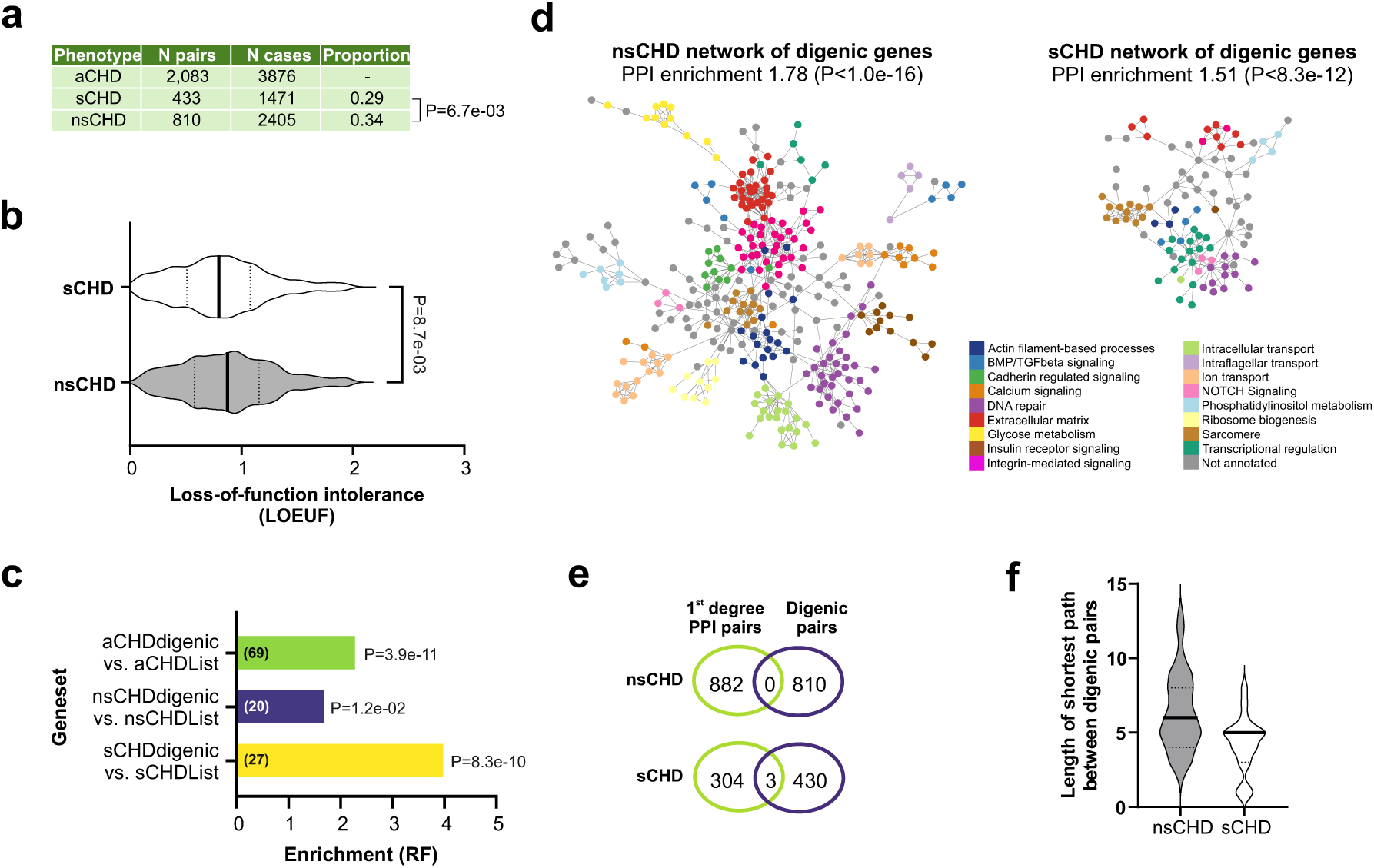
Case-control enrichment analysis at the digenic level using the *RareComb* framework. **(a)** Proportion of digenic pairs contributing to syndromic and non-syndromic CHD identified at FDR 1%. **(b)** Comparison of the distribution of LOEUF metric (at gene level) between syndromic and non-syndromic. The analysis was performed on genes observed in just one digenic pair. **(c)** Overlap of genes forming digenic interactions with known CHD genes. **(d)** Protein-protein interaction (PPI) network analysis of digenic genes for sCHD and nsCHD (PPI networks with annotated gene names are shown in **Supplemental Figure 10**). **(e)** Overlap between digenic pairs and first-degree interactors for sCHD and nsCHD in its respective PPI networks. **(f)** Length of the shortest path observed between genes forming a digenic pair in a PPI network.

The rate of a gene being observed in a digenic pair showed no correlation (r2 < 0.2) with its coding sequence (CDS) length (**Supplemental Figure 9a**). Thus, the digenic interactions implicated by our data does not appear to be biased by an increased mutation rate of larger genes.

A significantly lower LOEUF was observed in the genes forming digenic pairs contributing to syndromic CHD compared to non-syndromic CHD (**Figure 5b**). A similar pattern was observed by comparing the distribution of MOEUF (**Supplemental figure 9b**), whereas no significant differences were observed when analysing the observed/expected synonymous upper fraction ratio metric (SOEUF, **Supplemental figure 9c**). Thus, our data suggest that digenic pairs show similar correlation between loss-of-function intolerance and sCHD as we observed for single genes (**Figure 2b**). We hypothesized that if genes in the digenic gene-sets are causative of CHD in our patient cohort, we would expect that they are enriched for known CHD genes. To test this hypothesis, we calculated the overlap between genes in the digenic lists aCHD, sCHD and nsCHD and curated lists of genes known to cause CHD in patients^8^. We observed significant enrichment of known CHD disease genes in all three digenic lists (**Figure 5c**).

To investigate if digenic pairs interact at a systems level, we generated protein-protein interaction (PPI) networks using data from STRING^18^. Networks of 1,929 nodes/3,587 edges, 1,042 nodes/882 edges and 610 nodes/307 edges were generated for aCHD, nsCHD and sCHD digenic lists, respectively (**Figure 5d, Supplemental Figure 10**). Analysis of the networks showed that nsCHD and sCHD digenic gene-lists interact in physical networks with PPI enrichment values of 1.78 (*P* < 1.0 × 10^-16^) and 1.51 (*P* = 8.3 × 10^-12^), respectively.

To test if the genes in digenic pairs share the same function, we performed a simple overlap test to see how many digenic pairs show direct (1^st^ degree) interaction in the network. The results show that only very few pairs interact directly at the protein level (**Figure 5e**). Finally, to determine the degree of separation between digenic pairs in each network, we calculated the length of the shortest path between genes in each pair. This analysis showed that the median of the shortest path between digenic pairs is 5 and 6 for the sCHD and nsCHD networks, respectively (**Figure 5f, Supplemental Figure 11**).

In summary, our analyses suggest that the digenic pairs interact at systems level in complex PPI networks, with a high degree of separation between the genes in each digenic pair.

## DISCUSSION

In this study, we amassed 57,628 human exomes and conducted both a gene- and gene-set centred case-control burden analysis to increase our understanding of the genetic causes of CHD. After quality control at the sample and variant level, we provide a comprehensive CHD case-control cohort with unrelated and ancestry-matched individuals. Specifically, the availability of detailed phenotype data allowed us to explore the differences between syndromic and non-syndromic forms of CHD. By utilising gene-level constraint information^10^, we investigated the contribution and properties of loss-of-function and missense constraint variants independently for all CHD cases, as well as syndromic and non-syndromic CHD independently. Like earlier comparable studies^6,7^, our results revealed a higher contribution of LOF variants to CHD compared to missense variants, confirming that this type of variation represents the largest driver. Subsequently, the analysis of syndromic cases revealed a higher burden of LOF mutations when compared with the non-syndromic cohort. This effect was mainly a result of the contribution of genes with a higher intolerance to loss-of-function variations. This same pattern was also observed when analysing the genes based on missense constraint.

We next assessed the contribution to CHD at the gene level, by performing a gene-based case-control burden analysis. Our analysis revealed ten genes that reached genome-wide significant levels of association with CHD (*NSD1*^19^, *TAB2*^20^, *KAT6A*^21^, *PTPN11*^22^, *CTCF*^23^, *SMAD4*^24^, *FLT4*^25^, *NOTCH1*^26,27^, *BCOR*^28^ and *KMT2A*^29^). Previous studies have associated these genes as a cause of CHD, and our results confirm this association (**Table 1**). Furthermore, four candidate genes (*PBX1, SHOX2, KAT6B, HCAR1*) were found contributing to both syndromic and non-syndromic CHD at *FDR* 5%. To our knowledge, these genes have been either not previously associated with, or have only been briefly described in the context of CHD.

*PBX1* has been primarily associated with congenital abnormalities of the kidney and urinary tract (CAKUT)^30^; however, previous studies have reported isolated cases carrying *de novo* missense variations leading to syndromic CHD^30,31^. In line with these early reports, our analysis revealed a significant burden of missense constrained variants in *PBX1* in syndromic CHD patients (**Table 1**). It has also been demonstrated that deficiency of *Pbx1* impacts branchial arch artery patterning and results in the failure of cardiac outflow tract septation^32^. Interestingly, this gene was also found to be differentially expressed in Epicardium and Smooth muscle cells (**Supplemental Figure 12**). Together our findings suggest that *PBX1* contributes significantly to syndromic forms of CHD.

*SHOX2* was significantly enriched (at *FDR* 5%) in the nsCHD cohort, for missC variants (**Table 1**). Recent studies in animal models have demonstrated that the *Shox2* null mice are embryonic-lethal^33^. Cardiovascular defects identified in these mice included an abnormally low heartbeat rate, a severely hypoplastic Sinoatrial Node (SAN), hypoplastic or absent sinus valves^33^, and other atrial abnormalities (e.g., enlarged atrial chamber and thinner atrial wall). Subsequently, *SHOX2* has been described as playing a key role in developing the Sinoatrial Node^33,34^. In addition, *SHOX2* was identified as a significant DEG in atrial cardiomyocytes (**Supplemental Figure 12**), providing further supporting evidence of its role in heart development, most likely by regulating the activity of *NXK2-5*^33,35^ and *TBX5*^36^. These results imply that *SHOX2* is a plausible novel non-syndromic CHD gene.

Truncating variants in the Lysine Acetyltransferase 6B gene (*KAT6B*) have been associated with Say–Barber–Biesecker–Young–Simpson Syndrome (SBBYSS, OMIM 603736) and Genitopatellar Syndrome (GTPTS, OMIM 606170). Heart defects have been reported as part of the phenotypic spectrum of SBBYSS^37^. In a recent study of 32 individuals with *KAT6B* disorder, 47% showed cardiovascular anomalies, mainly atrial septal defects, ventricular septal defects, and patent ductus arteriosus^38^. Our results have identified that *KAT6B* was differentially expressed in the cluster of atrial cardiomyocytes cells (**Supplemental Figure 12**), which suggests a possible role in the early cardiac development program. Our analysis extends previous findings associating loss-of-function variations in *KAT6B* to sCHD.

The Hydroxycarboxylic Acid Receptor 1 (*HCAR1*) does not appear to have been associated with CHD thus far, however our findings suggest this gene may be a novel candidate CHD gene. It was not differentially expressed in any of the cardiac-specific cell clusters analysed.

By meta-analysing the genomic data with heart single-cell transcriptomic data, we investigated the pattern of expression of DEGs for aCHD, sCHD and nsCHD in a range of cardiac-specific cells. Using Gene Ontology enrichment as a complementary analysis, we identified key gene markers and biological processes associated with CHD. Unlike previous studies^7,39^, which focused on whole heart bulk-RNA sequencing data, the use of transcriptomic data at a single-cell resolution allowed the analysis of candidate gene expression patterns in specific cardiac cell clusters important for early cardiac development. Our analysis highlighted distinct cardiac cell clusters contributing to sCHD and nsCHD. In addition, we demonstrated that missense constrained variants could have a similar functional impact compared to loss-of-function variants, although to a lesser degree. For instance, the significant enrichment of sCHD in cardiac neural crest cells (cNCCs) suggests a broader contribution of patients affected by syndromic occurrences, not limited to heart development only. Perturbations in the cNCCs migration process can lead to a wide spectrum of human cardio-craniofacial syndromes, including DiGeorge Syndrome (22q11.2 Deletion Syndrome, OMIM 188400) and CHARGE (OMIM 214800). The enrichment observed in capillary endothelium and pericyte cells in nsCHD, associated with the vasculogenesis process, suggests that the phenotypic occurrence in these patients is limited to the cardiovascular system rather than affecting a broader spectrum of cells. Whilst the results are promising, they are limited because the currently available human heart single-cell map^9^ is incomplete (e.g., only a few early developmental time points). Therefore, future studies integrating mouse and human single-cell heart and whole-embryo data are warranted.

Contrasting with the study of monogenic causes of CHD, oligogenic factors underlying the disease have been explored to a lesser extent. We took advantage of a newly developed method to study the contribution of digenic interactions (the simplest form of oligogenic) to CHD, in a case-control setting^17^. Interestingly, we observed a higher proportion of digenic interactions contributing to non-syndromic compared to syndromic CHD. These results contrast with those observed at the gene level, where 16 out of 21 genes (∼76%) found significant at *FDR* 10% (**Table 1**) were associated with syndromic CHD. These findings imply that sCHD is more likely to have a monogenic aetiology, and oligogenic interactions may be a more important component in the development of non-syndromic forms of CHD.

Functional annotation of subclusters in the networks, generated from nsCHD and sCHD digenic pairs, indicate that the identified digenic pairs encode proteins involved in transcriptional regulation, signalling pathways (e.g., BMP/TGF beta signalling and NOTCH signalling) and tissue structures (e.g. sarcomere and extracellular matrix) which are important in heart development^40–42^.

Our network analyses suggest that rather than interacting within the same subcluster, the digenic pairs interact at systems level, with a high degree of separation between the genes in each pair, thus supporting previous results which suggest that CHD risk factors converge in higher-order developmental networks^43^.

In summary, we analysed ∼57,000 exomes, and complemented this with transcriptomic data at single-cell resolution. The findings have strengthened the association of previously described genes with CHD, identified novel candidate genes, and provide a deeper understanding of the pathophysiological mechanisms underlying CHD at gene and digenic level and the potential different aetiologies between syndromic and non-syndromic CHD.

## METHODS

### Cohort description

To create a comprehensive CHD case-control cohort, exome sequencing data from multiple individuals was combined in a unique reference dataset. CHD cases were mainly sequenced as part of an initiative from the German Competence Network for Congenital Heart Defects, the Deciphering Developmental Disorder (DDD) project and the University of Nottingham (UK); controls were sequenced as part of the UK Biobank (UKBB). Samples from the UKBB dataset with phenotype description labelled as Schizophrenia (SCZ), bipolar disorder (BP) or developmental delay (DD) were excluded from the analysis. Accordingly, a small fraction of samples in the UKBB cohort (127 samples), labelled as CHD cases, were included in the analysis. In total, we assembled an exome dataset consisting of 57,628 samples (4,747 CHD cases and 52,881 controls).

### Alignment, quality control and variant annotation

The assembled dataset was processed and harmonized using the same alignment (BWA v0.3), calling (GATK v4.0), annotation (VEP v95) and quality control (Hail v0.2) pipelines. **Supplemental Information 1** describes extensively the implementation and results of these methods.

### Defining a set of loss-of-function and missense constraint variants

We enriched the dataset for high-confidence loss-of-function (hcLOF) variants and missense constrained (missC) variants. hcLOF variants were annotated as indicated by the LOFTEE tool (https://github.com/konradjk/loftee) with its default parameters and included stop-gained, essential splice and frameshift variants. To define a set of missC variants, we evaluated four state-of-art pathogenicity prediction scores: CADD^44^, MPC^45^, REVEL^46^ and MVP^47^. Specifically, the performance of these scores was assessed by classifying benign and pathogenic missense variants (accessed through the ClinVar database, https://www.ncbi.nlm.nih.gov/clinvar) in the context of known CHD genes. In brief, receiver operating characteristic (ROC) analysis was conducted for benign and pathogenic variants within known CHD genes. The analysis was further stratified by splitting the gene set into LOF constraint (LOEUF < 0.35) and LOF non-constraint (LOEUF >= 0.35) genes. A score was defined as a ’good predictor’ if achieved an area-under-curve (AUC) > 90% in both evaluated scenarios. Three of these scores (CADD, REVEL and MVP) met this criterion. A missense variant was defined as missC if it was predicted as likely deleterious by at least two of these scores based on the optimal threshold suggested by the ROC analysis (**Supplemental Figure 2**).

### Defining rare variants

Variants were filtered based on the cohort-specific allelic frequency (’internal’ AF) as well as using external datasets. A variant was defined as rare if AF was lower than 0.001 (MAF < 0.001) in the gnomAD database^10^ (both exomes v2.1.1 and genomes v3.0.0), the RUMC cohort^48^, as well as AFs from an *in-house* German exome sequencing cohort.

### Gene-set enrichment analysis

*Generation of gene sets.* Gene set-level association analysis was performed to assess whether an excess of the possible pathogenic variants was enriched for a particular category of genes (as described below). This procedure was executed for the following gene sets:

a. LOEUF gene bins: Constraint loss-of-function (LOF) metrics per protein-coding genes were accessed through gnomAD resource^10^. Genes were ranked by their observed/expected LOF mutation ratio upper fraction (termed LOUEF), and ten bins with an equal number of genes (∼1,900 genes per bin) were defined. Lower values of LOEUF (e.g., bins 1 and 2) denote most LOF-constrained genes.
b. MOEUF gene bins: Similar as described above for LOEUF genes, but genes were binned based on their observed/expected missense mutation ratio upper fraction (termed MOEUF).
c. Differentially expressed genes (DEGs) in cardiac-specific cells: DEGs identified in 15 distinct cardiac cell clusters reported by Asp *et al*^9^. In brief, genes were determined as significantly differentially expressed in a particular cardiac cell cluster if the averaged log-fold change (logFC) > 0 (upregulated) at FDR 1%.

*Gene set-based association analysis.* For each sample within the filtered dataset, we generate a Minimal Allele Count (MAC) metric by aggregating high confidence Genotypes (DP >= 10, GQ >= 20 and allelic balance heterozygous > 0.2) across the genes within the gene set. Then, a burden logistic regression test was performed using CHD case/control status as response and the five first ancestry principal component and sex as covariates using the Hail function *hl.logistic_regression_rows*. The analysis was stratified at the sample and variant level. At the sample level, the data was divided based on the syndromic status; three categories were tested: aCHD (all CHD cases vs control), nsCHD (non-syndromic CHD cases vs control) and sCHD (syndromic CHD cases vs control). At variant level, three different groups were evaluated based on the predicted severity of the variants: hcLOF (most severe), missC and synonymous. The synonymous variant set was used as a negative control set at the variant level to evaluate for potential artefacts. The odds ratio (exp (beta coefficient)), 95% confidence interval and *p-value* metrics were used to evaluate significant enrichment.

### Gene-based burden testing

We performed case-control gene-centred burden test analysis to assess genes with significant association with CHD. Fisher Exact test was performed independently for rare (MAF < 0.001) hcLOF and missC variants. To define the significant study-wide *p-value*, the minimal *p-value* (*P*) per gene between these two categories was chosen. The analysis was further stratified by syndromic status to assess the distinct contribution of these categories to CHD. A gene was defined as genome-wide significant if it reached a Bonferroni corrected *P* < 0.05 and suggested significant if *FDR* < 5%. In addition, the set of synonymous variants was used as a negative control set since no difference between cases/control is expected on this set of variations (quantile-quantile plots, **Supplemental Figure 3**).

### Expression analysis using bulk RNAseq data

A publicly available human transcriptomic dataset previously described by Cardoso-Moreira *et al*^15^ was used to complement this study. To assess the gene expression levels in the heart, kidney, brain, and liver; the RPKM matrix hosted in ArrayExpress (E-MTAB-6814) was used. Gene expression levels were averaged among samples in the early developmental stages (4-8 weeks-post-conception). Percentile rank per gene was computed based on the mean expression.

### Gene Ontology enrichment analysis

The R-package *Enrichr* (with the *Biological_Process_2018* database) was used to perform Gene Ontology (GO) enrichment analysis. The analysis was conducted on the differentially expressed genes (DEGs) in cardiac-specific cell clusters, which also showed unadjusted *P* < 0.01 (Fisher Exact test) from the case-control burden analysis. The evaluated DEGs were previously reported by Asp *et al*^9^ with no additional processing. GO terms with only one overlapping gene were filtered out. A biological process term was considered significant if *FDR* < 1% as reported by the Enrichr tool^15^.

### Digenic analysis

The digenic analysis was performed using the R-package RareComb^17^. RareComb combines inferences statistics with an a priori algorithm to elucidate digenic/oligogenic combinations that are enriched for rare genetic variants. Specifically, we implemented the test *‘enrichment_depletion’*, to access digenic pairs significantly enriched with rare (MAF < 0.001) hcLOF and/or missC variations in CHD cases compared to controls (depleted). The analysis was further stratified by syndromic status (aCHD, sCHD or nsCHD vs. controls). Digenic pairs were defined as significant if *FDR* < 1%.

Enrichment of known CHD genes was determined by calculating overlap between gene lists. Significant overlap was calculated using hypergeometric statistics. A representation factor was calculated as the number of overlapping genes, divided by the expected number of overlapping genes drawn from two independent groups: RF=x/((n*D)/N), where x=number of overlapping genes, n=genes in group 1, D=genes in group 2, N=protein-coding genes in genome (20,000).

Protein-protein interactions (PPIs) were obtained from STRING v.11.5 using genes in the digenic pairs to query the database. The following parameters were used; network type: physical subnetwork, active interaction sources: experiments, databases (text mining data excluded), minimum required interaction score: 0.400 (medium confidence). PPIs were visualized as a network using CytoScape v3.9.1; nodes represent proteins and edges represent interactions between these proteins. PPI enrichment was analysed using the Analysis tool available in the online version of STRING (https://string-db.org). PPI enrichment was calculated as (observed number of edges) / (expected number of edges) and PPI enrichment P-values were obtained directly from STRING. Length of the shortest path between genes in each digenic pairs, within the nsCHD and sCHD networks was calculated using PesCa^49^ v3.0.8.

## Data availability

The CRAM-level data from CHD patients used in this study can be accessed under the following accession codes (European Genome-phenome Archive): EGAD00001002200, EGAD00001000796, EGAD00001000797, EGAD00001000800, EGAS00001000544, EGAS00001000775, EGAS00001000762. UK Biobank 50K WES dataset freeze was accessed under the application number 44165.

## Code availability

Pipelines for sample/variant quality control (QC), annotation, and burden testing are available on GitHub: https://github.com/enriquea/wes_chd_ukbb.

## Acknowledgements

This research was conducted using the UKBB Resource under application number 44165. We used data from the Deciphering Developmental Disorders (DDD) study. The DDD study presents independent research commissioned by the Health Innovation Challenge Fund, a parallel funding partnership between the Wellcome Trust and the UK Department of Health, and the Wellcome Trust Sanger Institute. The views expressed in this publication are those of the author(s) and not necessarily those of the Wellcome Trust or the UK Department of Health. The authors wish to thank Prof. Matthew Hurles (Sanger Institute, UK) for his significant contribution to this study. We thank the KinderHerzen e. V. for providing research funding for this study. We thank Prof. Dr. Christian Gilissen (RadboudUMC), Prof. Dr. Peter Krawitz (University Boon), and collaborators from the Universitaetsklinikum Tuebingen (Prof. Dr. Stephan Ossowski, Prof. Dr. Olaf Horst Rieß, Prof. Dr. Tobias Haack), for providing us with Central European allele frequencies. This work was partly funded by PROCEED project ERA PerMED joint Translational Call Initiative (DLR Funding reference number: 01KU1919).

## Supplemental figures

**Supplemental Figure 1.**
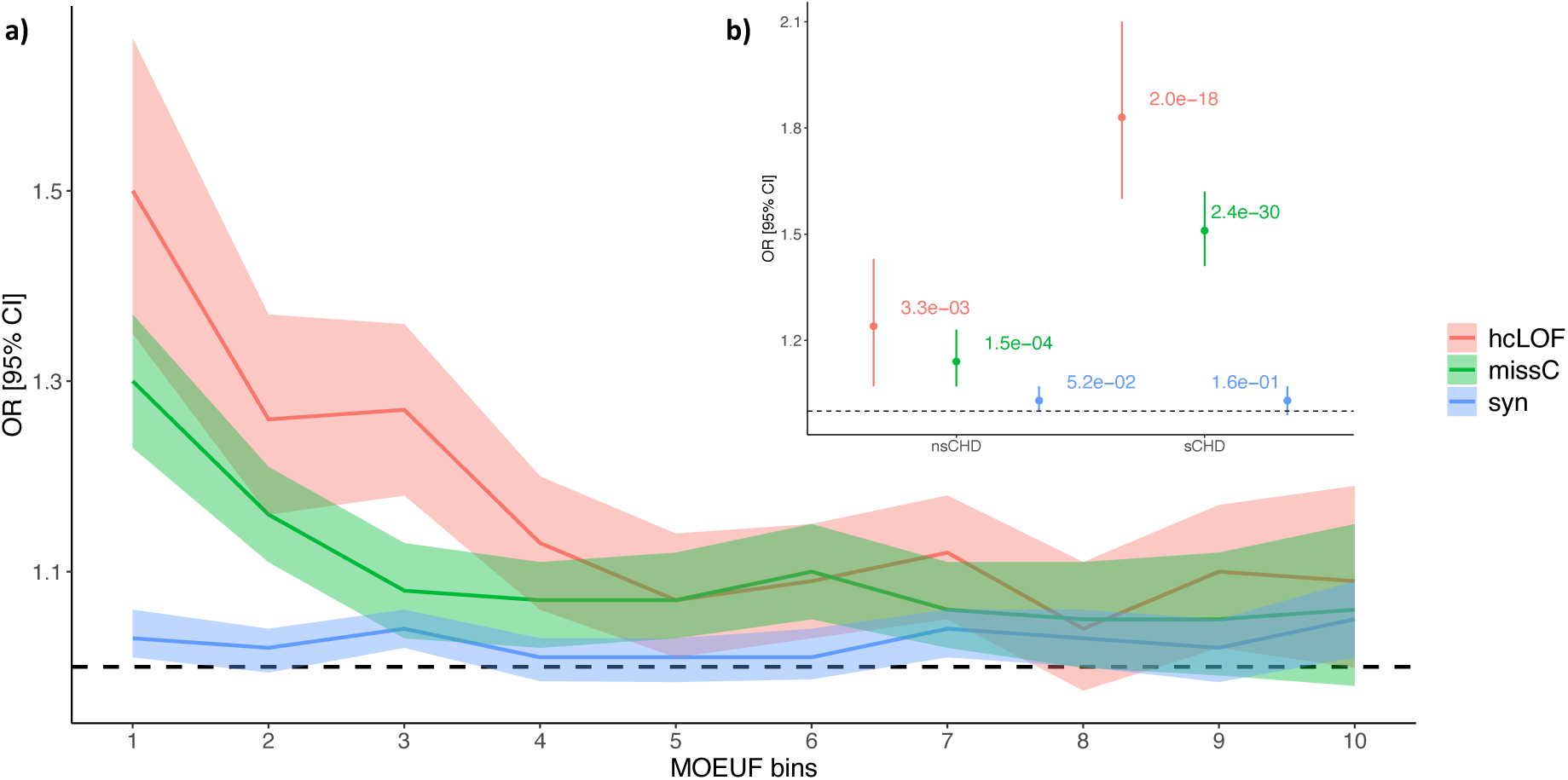
Enrichment analysis across the missense constraint gene spectrum. Protein-coding genes were binned based on the MOEUF metric as proposed by gnomAD. Every bin contains ∼1,900 genes. Top bins (1, 2) contain the genes with the highest intolerance to missense variation. **a)** Enrichment analysis per bin for aCHD vs controls are shown. **b)** Enrichment analysis stratified by syndromic status (sCHD and nsCHD) vs controls in the top constraint MOEUF bin (1). The x-axis indicates the constraint bins; the y-axis shows the Odd Ratios (OR) and the 95% confidence interval. hcLOF: high-confidence loss-of-function variants; missC: missense constrained variants; syn: synonymous variants.

**Supplemental Figure 2.**
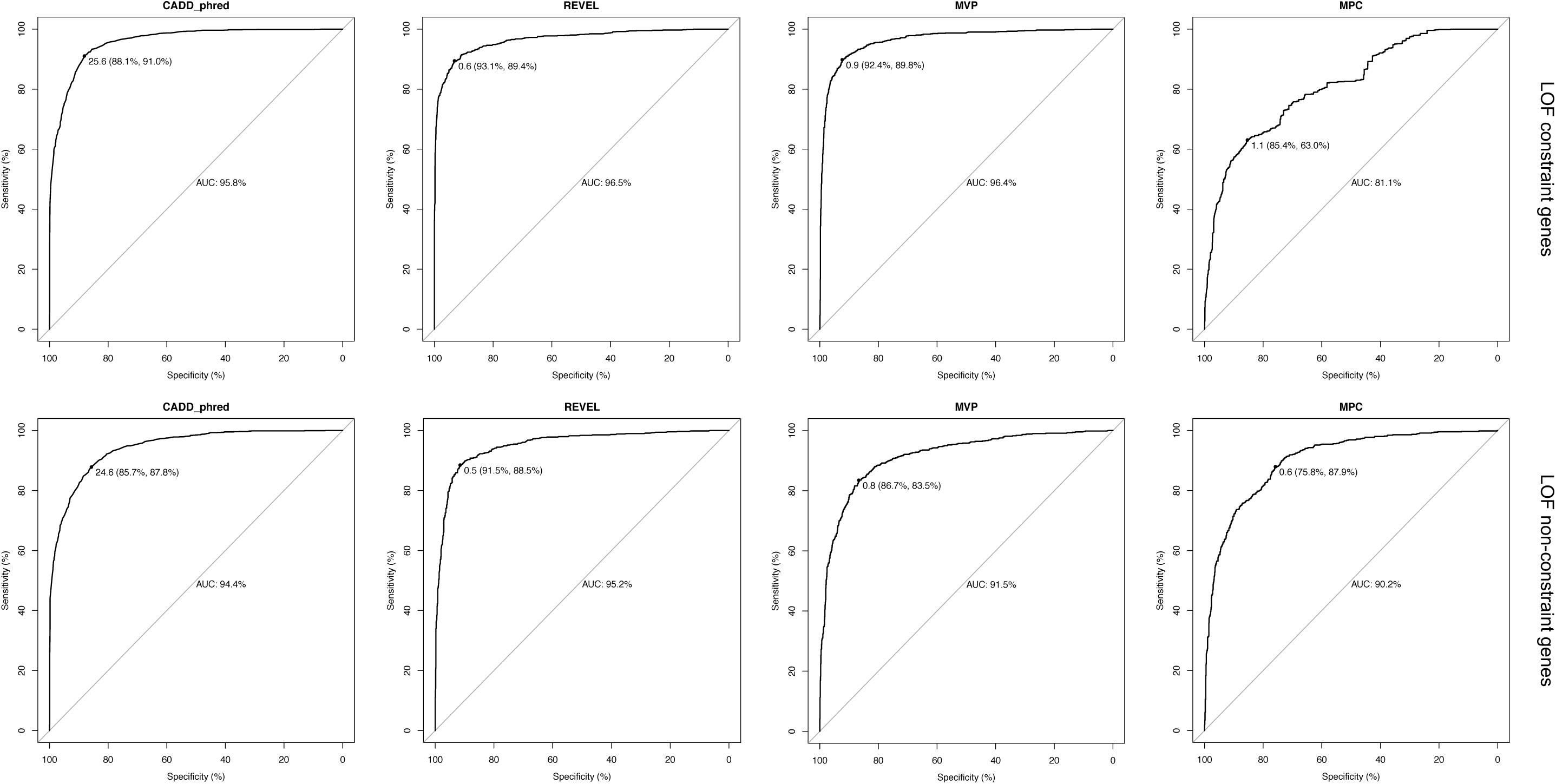
ROC analysis of pathogenicity prediction scores (CADD, REVEL, MVP and MPC). The analysis was performed on a balanced set of benign (true negative) and likely pathogenic (true positive) variants from the ClinVar database within known CHD genes. The top panels show the results for LOF constraint genes (LOUEF < 0.35). The bottom panels show the results for LOF non-constraint genes (LOUEF >= 0.35).

**Supplemental Figure 3.**
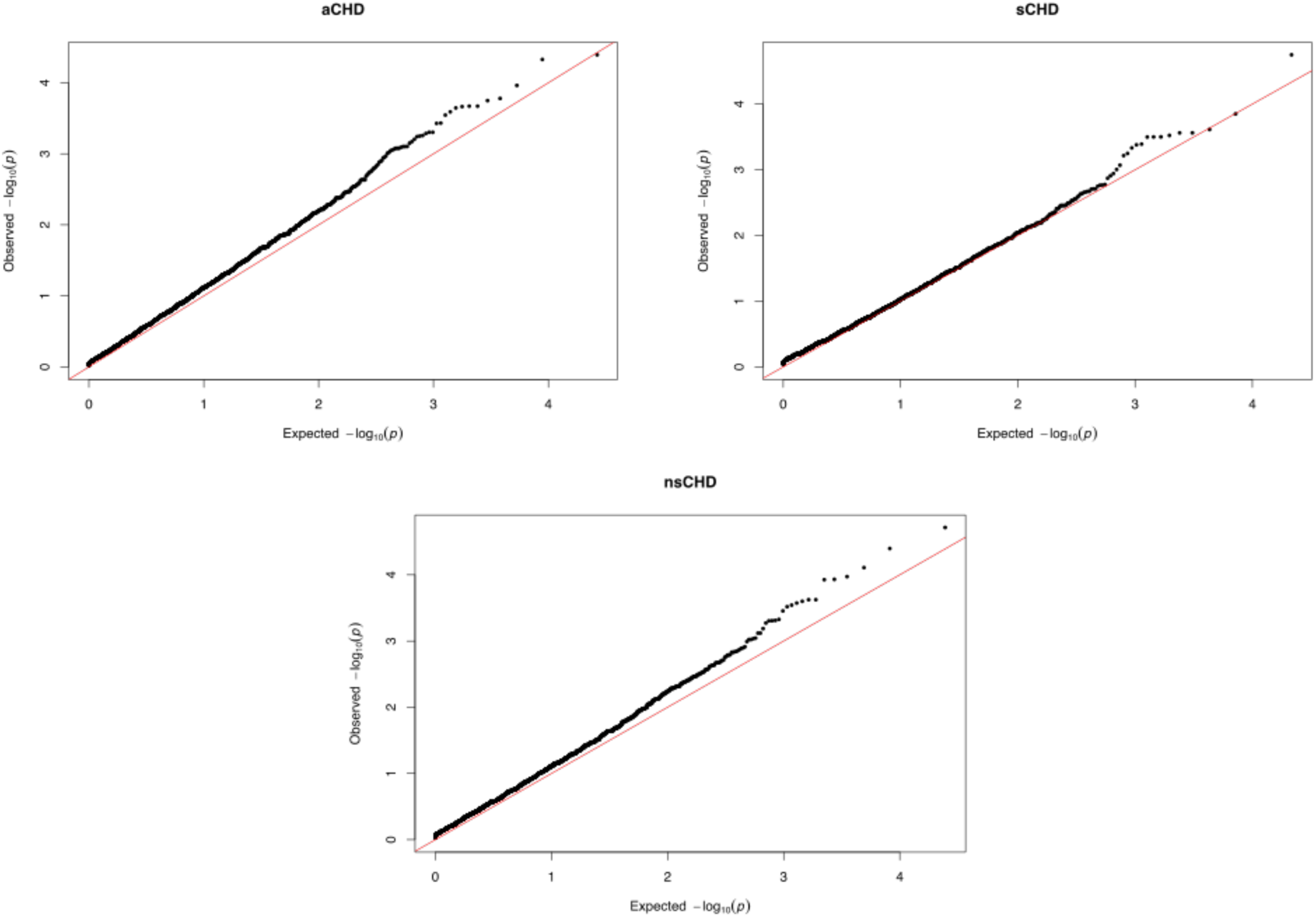
Quantile-quantile plots. Expected vs observed p-values for synonymous variants stratified by syndromic status (MAF 0.1%). Q-Q plots for aCHD, sCHD and nsCHD vs controls are shown.

**Supplemental Figure 4.**
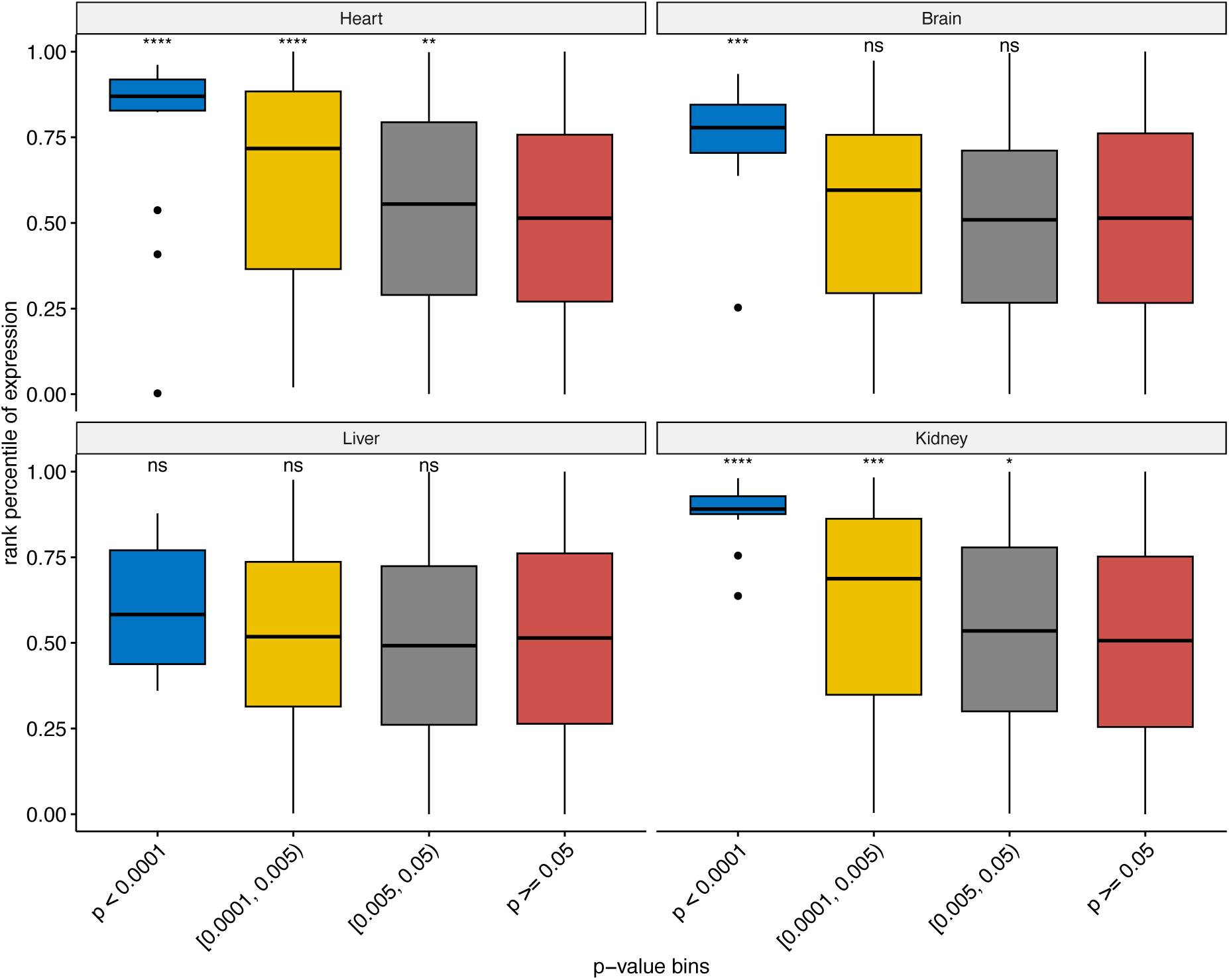
Expression pattern of CHD genes in different tissues (Heart, Brain, Liver and Kidney). X-axis denotes gene p-value bins. P-value refers to the minimal p-value (*P*) observed in the gene-based enrichment analysis for rare hcLOF and missC variants. The gene association analysis was performed by comparing all CHD probands (aCHD) vs controls. Y-axis denotes tissue-specific percentile rank of mean expression. Averaged expression was computed for samples between 4-8 weeks-post-conception (developmental stage). More significant genes (blue box) in the CHD case-control analysis showed the higher expression rank (e.g., Heart, Brain, and Kidney). Mean comparisons between bins were computed using the Wilcoxon test (alternative: greater; reference group (red box): genes with *P* > 0.05 in the case-control analysis). ns: p > 0.05; *: p <= 0.05; **: p <= 0.01; ***: p <= 0.001; ****: p <= 0.0001.

**Supplemental Figure 5.**
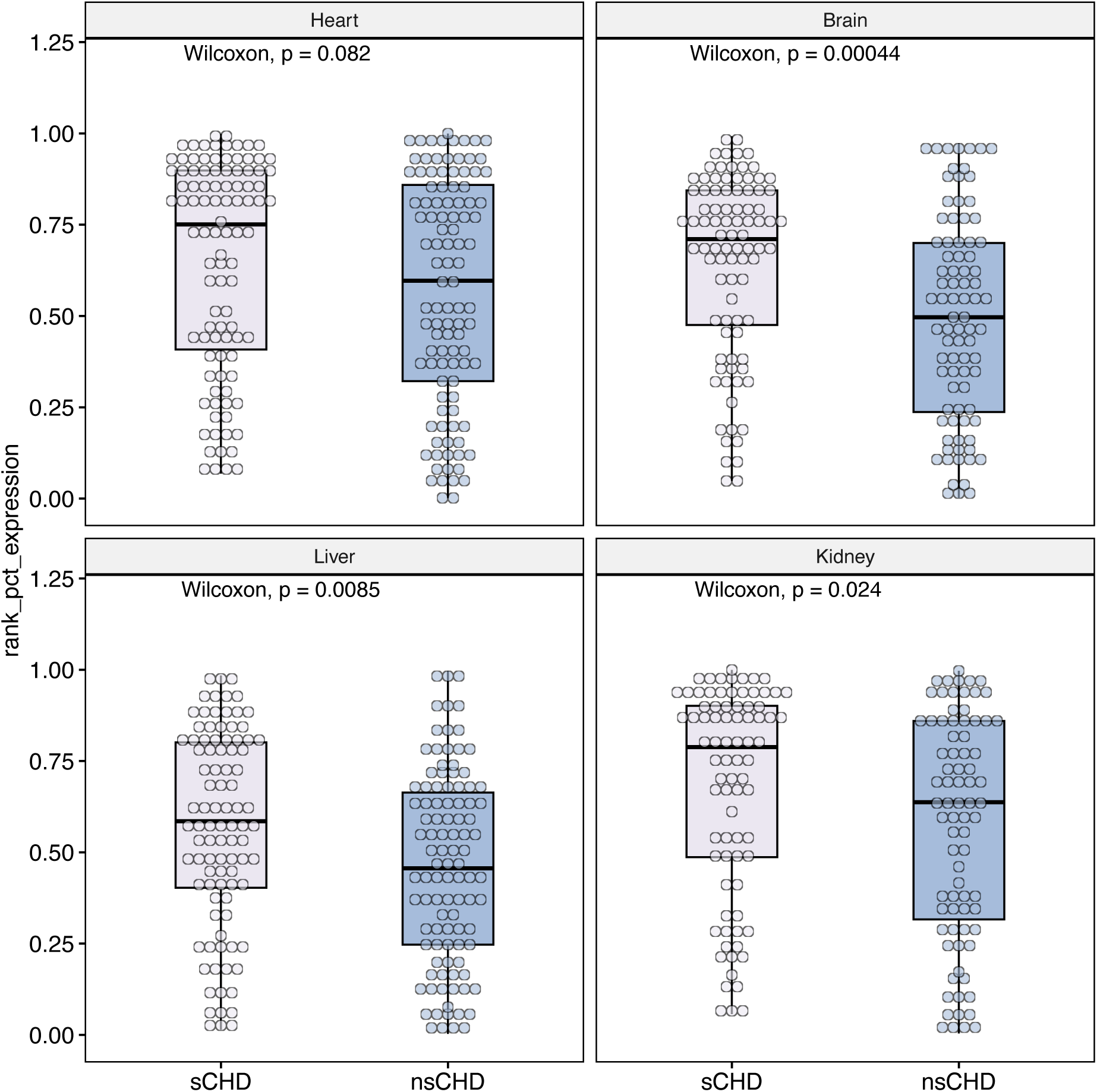
Tissue-specific expression pattern of CHD genes identified in the case-control analysis, stratified by syndromic status (syndromic (sCHD) and non-syndromic (nsCHD) vs controls). Only genes with unadjusted *P* < 0.005 in the case-control analysis are included. X-axis denotes the probands used in the case-control analysis (sCHD or nsCHD vs controls). Y-axis denotes tissue-specific percentile rank of mean expression. Averaged expression was computed among samples between 4-8 weeks-post-conception (developmental stage). Mean comparisons between groups were computed using the Wilcoxon test (two-sided). No significant difference was observed in the Heart for sCHD and nsCHD genes (*P* > 0.05), compared to other tissues (e.g. brain, liver and kidney, *P* < 0.05).

**Supplemental Figure 6.**
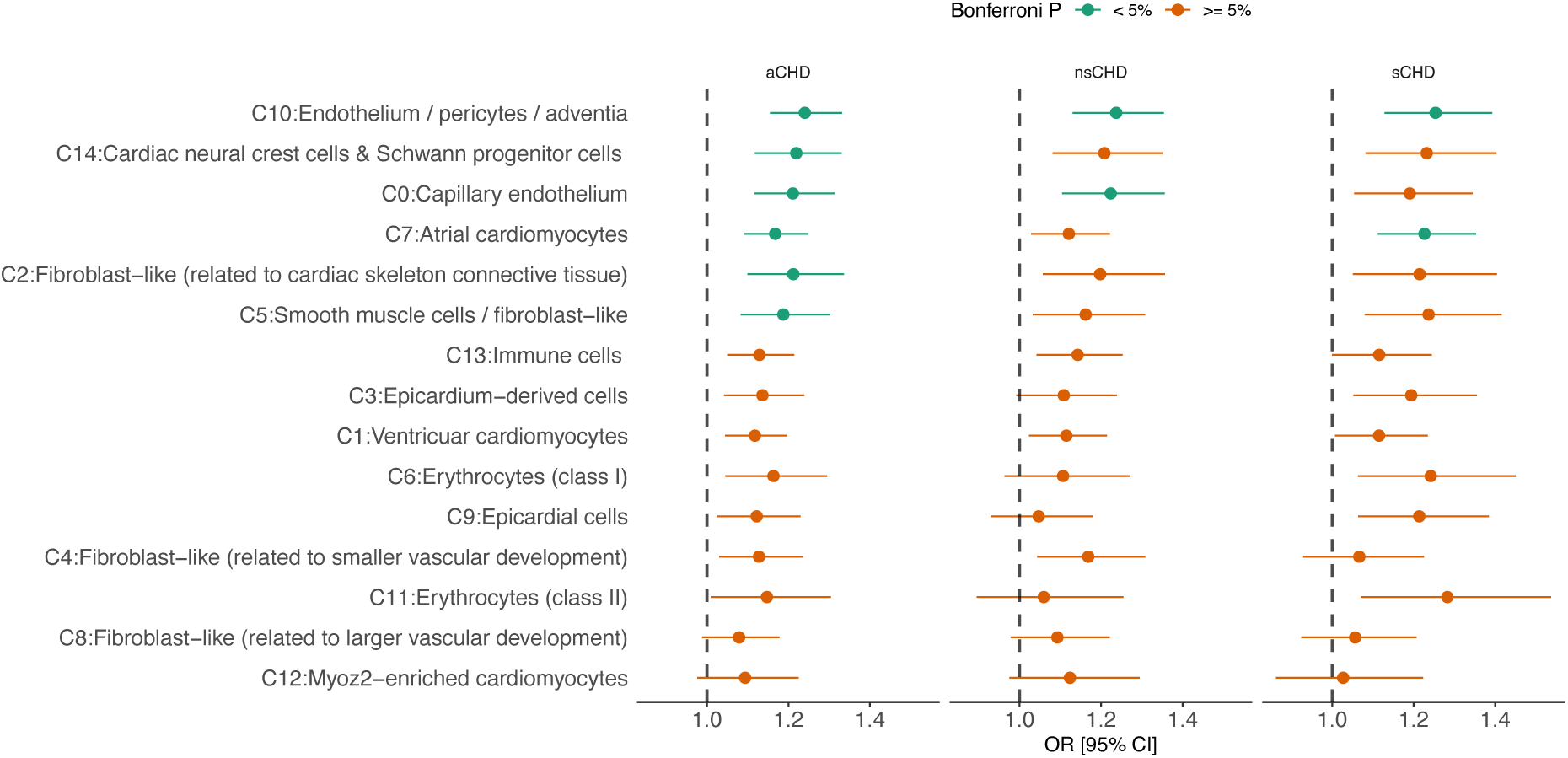
Logistic regression-based enrichment analysis of differentially expressed genes (DEGs) in cardiac-specific cell clusters for missense constrained variants (missC). The analysis was stratified by syndromic status (aCHD, sCHD and nsCHD). The x-axis denotes the Odds Ratio (OR) and the 95% confidence interval. *P-values* were adjusted using the Bonferroni method (0.05 / 45 tests) to assess for significant enrichment.

**Supplemental Figure 7.**
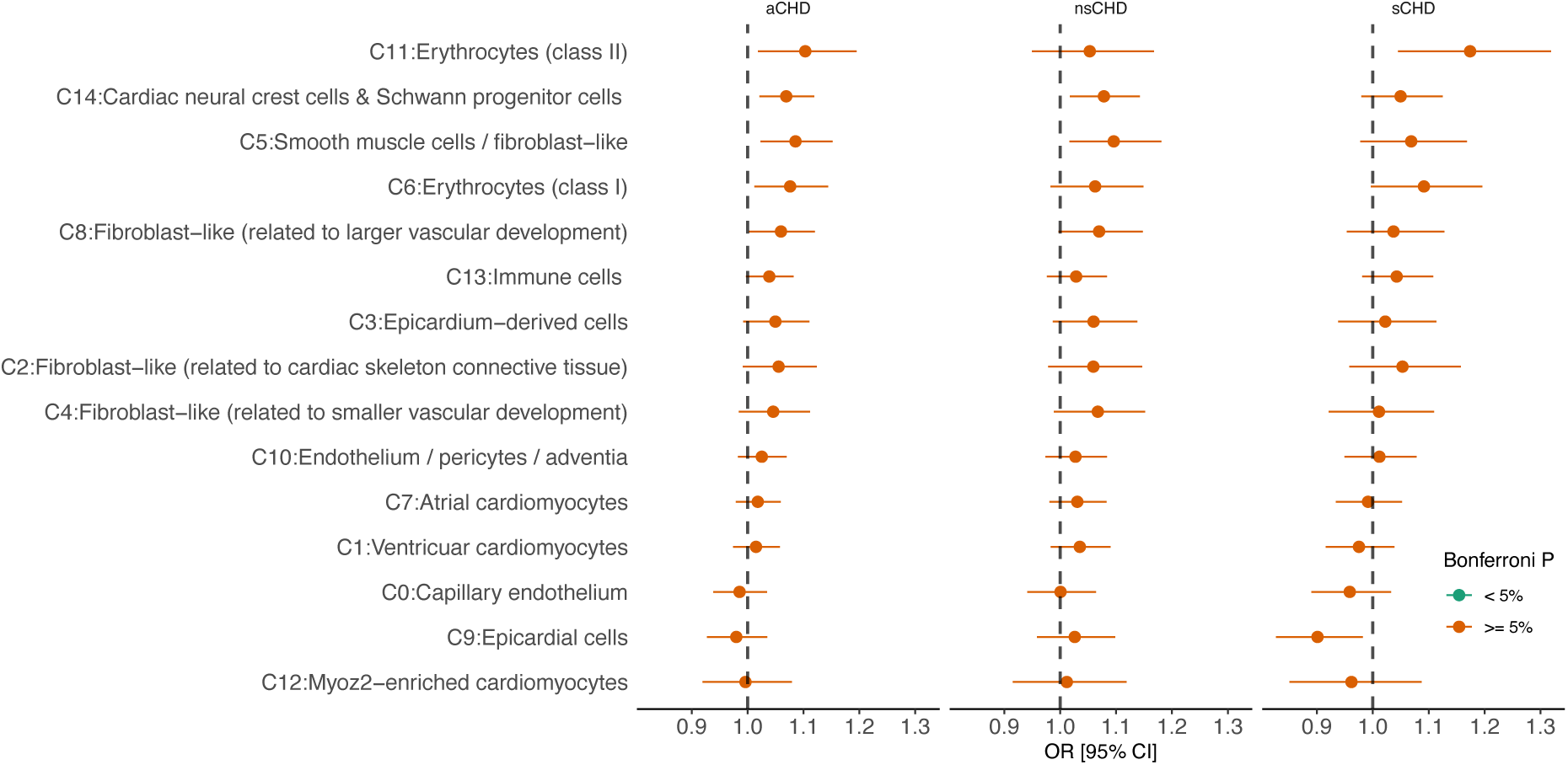
Logistic regression-based enrichment analysis of differentially expressed genes (DEGs) in cardiac-specific cell clusters for synonymous variants. The analysis was stratified by syndromic status (aCHD, sCHD and nsCHD). The x-axis denotes the Odds Ratio (OR) and the 95% confidence interval. *P-values* were adjusted using the Bonferroni method (0.05 / 45 tests) to assess for significant enrichment.

**Supplemental Figure 8.**
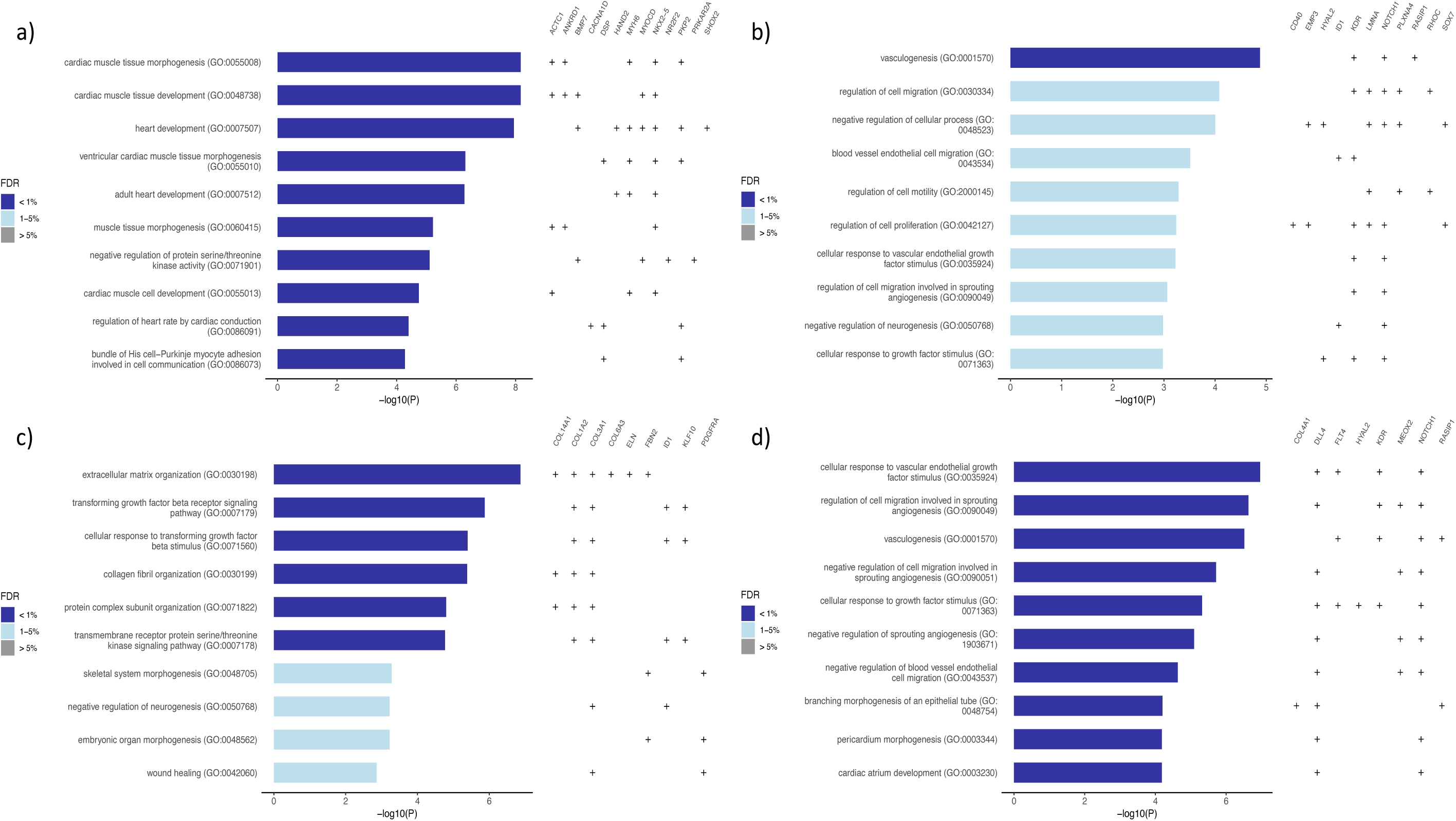
Gene Ontology (GO) enrichment analysis of differentially expressed genes (DEGs) in cardiac-specific cells with unadjusted *P* < 0.01 in the case-control burden analysis. a) C7: atrial cardiomyocytes cells, b) C0: capillary endothelium, c) C5: smooth muscle cells and d) C10: endothelium and pericytes cells. Only clusters with at least one GO term with *FDR* < 1% are shown. For every GO term, the overlapping DE genes (+) are shown.

**Supplemental Figure 9.**
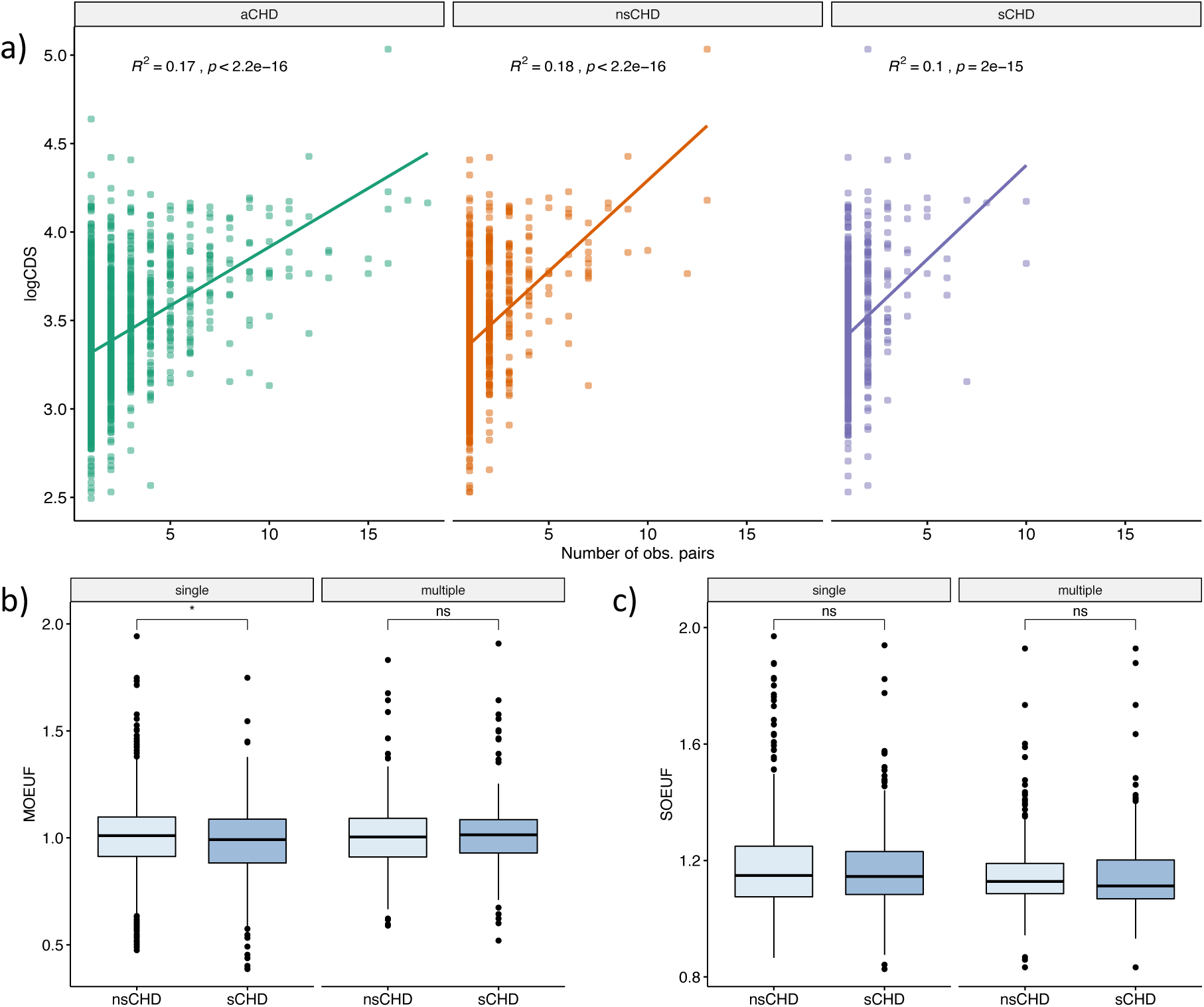
a) Correlation between the frequency at which a gene is observed forming multiple digenic pairs (x-axis) vs. its log-transformed CDS length (y-axis). The correlation analysis was stratified by syndromic status (aCHD, sCHD and nsCHD). b) Comparison of the distribution of missense observed/expected ratio upper fraction (MOEUF) metric (at gene level) between syndromic and non-syndromic. The analysis is stratified further into single-genes (i.e., genes observed in just one digenic pairs) or multiple-genes (i.e., genes that appears two or more times forming digenic pairs). c) Same as (b), but for the synonymous observed/expected ratio upper fraction (SOEUF).

**Supplemental Figure 10.**
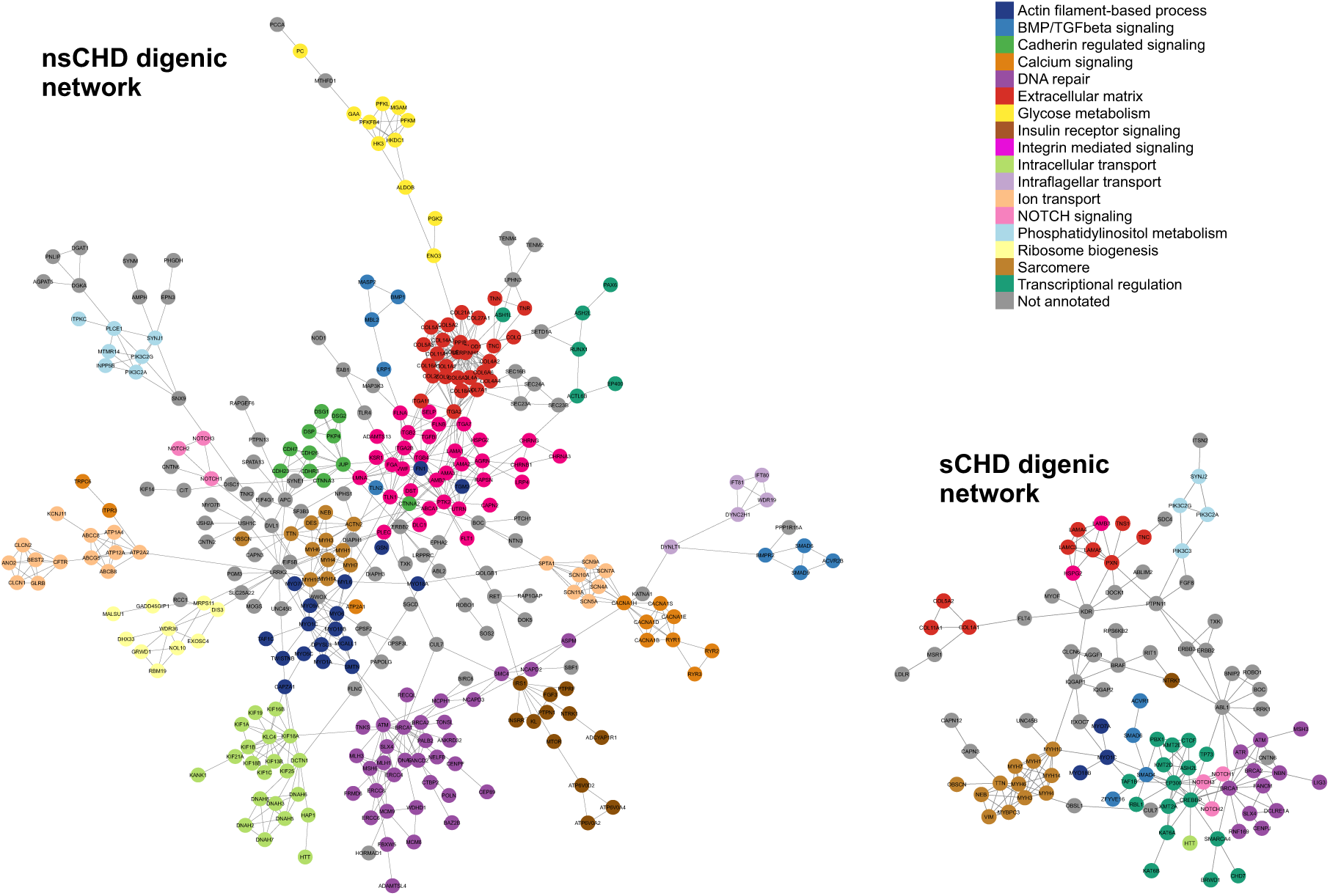
Protein-protein interaction network for syndromic and non-syndromic CHD digenic genes. Nodes are labeled with the corresponding gene name and annotated with the specific biological process.

**Supplemental Figure 11.**
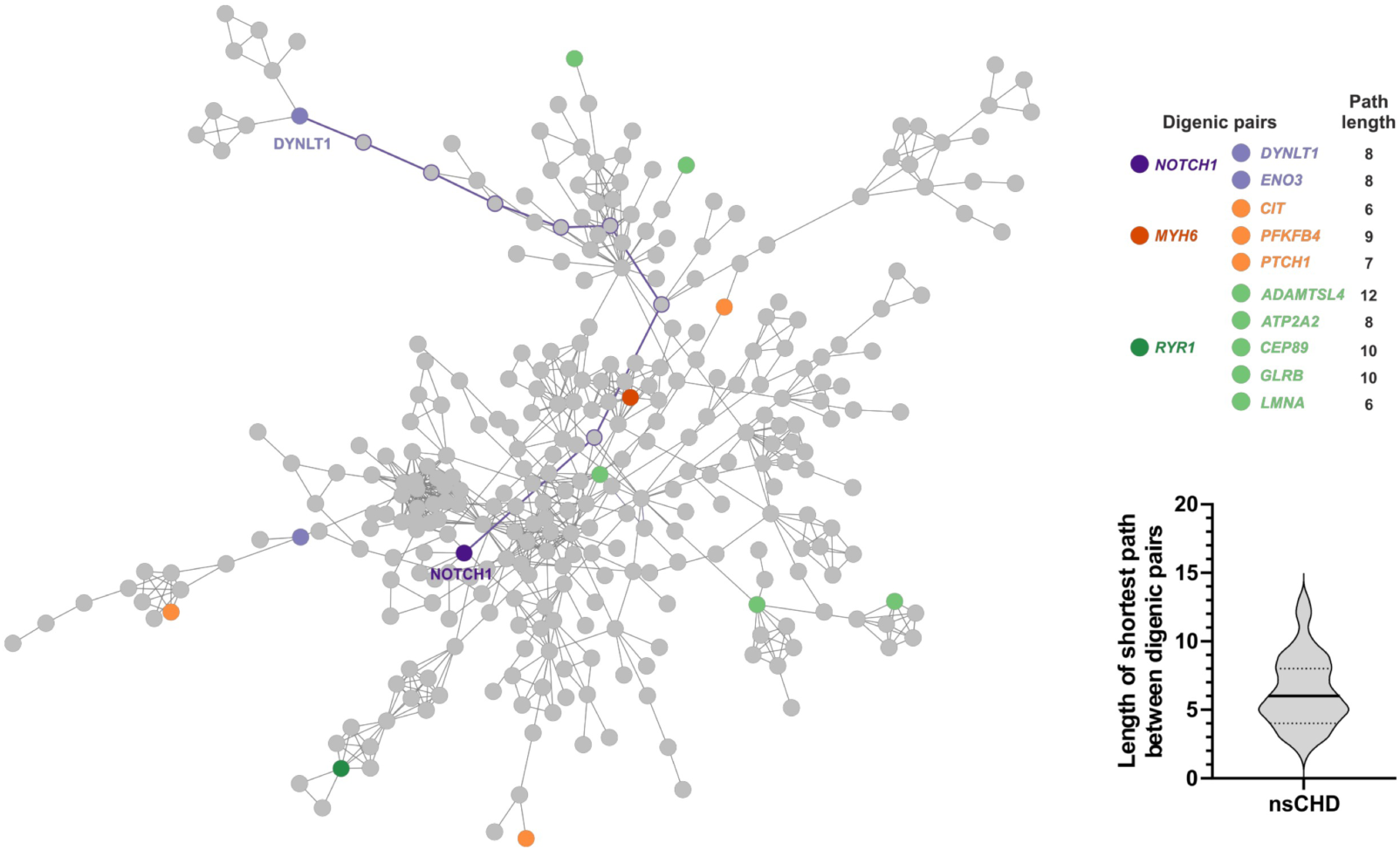
Digenic pairs are scattered across protein-protein interaction (PPI) network. Examples of non-syndromic digenic pairs and its shortest paths are highlighted in the network. The median of the shortest path between non-syndromic digenic pairs was six (violin plot).

**Supplemental Figure 12.**
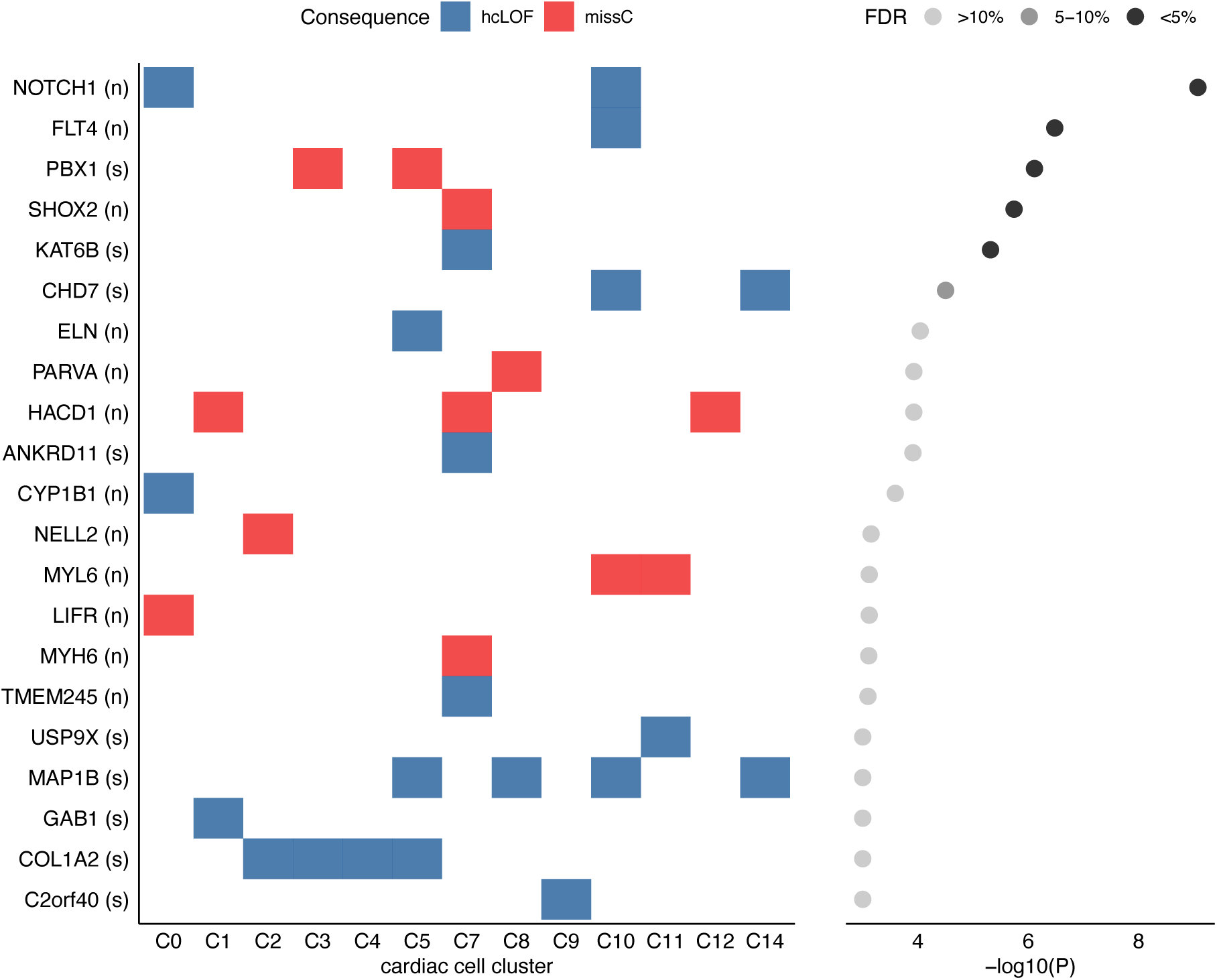
Top enriched genes (unadjusted *P* < 0.001, case-control Fisher Exact test) found differentially expressed in at least one cardiac-specific cell cluster. The left plot shows the gene/cluster overlap and highlights the variant category with the highest enrichment (blue: hcLOF, red: missC). The x-axis denotes de cardiac clusters; the y-axis indicates the genes and the CHD category analysed (s: sCHD, n: nsCHD). The right plot shows the log-transformed *P* (x-axis) and the *FDR* significant level per gene. Six genes showed *FDR* < 10%: *NOTCH1, FLT4, PBX1, SHOX2, KAT6B* and *CHD7*. C0: Capillary endothelium, C1: Ventricular cardiomyocytes, C2: Fibroblast-like (related to cardiac skeleton connective tissue), C3: Epicardium-derived cells, C4: Fibroblast-like (related to smaller vascular development), C5: Smooth muscle cells, C7: Atrial cardiomyocytes, C8: Fibroblast-like (related to larger vascular development), C9: Epicardial cells, C10: Endothelium/pericytes/adventia, C11: Erythrocytes (class II), C12: Myoz2-enriched cardiomyocytes, C14: Cardiac neural crest cells & Schwann progenitor cells.

