## Supplemental Information 1 for "Assessing the contribution of rare variants to congenital heart disease through a large-scale case-control exome study"

### Supplemental Information 1 (Extended methods)

### Alignment and variant calling

CRAM-level data for all previously and newly sequenced samples were realigned to the human genome build GRCh38 using the BWA tool (version 0.7). Variants were jointly called using the Genome Analysis Toolkit (GATK, version 4.1), following the Broad Institute best-practice guidelines for germline single nucleotide variants (SNVs) and short insertions/deletions (indels). Briefly, HaplotypeCaller was used in GVCF mode to process samples individually, such that every position in the genome was assigned with a likelihood of being or not being a variant. The GenomicsDB (https://github.com/Intel-HLS/GenomicsDB) tool was used then to import and merge the per-sample GVCF genotype data. Samples were then jointly genotyped for high confidence alleles using the GenotypeGVCFs tool. The Variant Quality Score Recalibration (VQSR) in GATK was applied independently for SNVs and indels to assess variant call accuracy. The complete process was executed using standard pipelines from the Human Genetics Informatics (HGI) unit at the Wellcome Trust Sanger Institute (WTSI).

To perform scalable downstream analysis of the sequencing data, the multi-sample cohort-VCF generated from the previous step was imported into Hail 0.2 (https://hail.is), a python-like library for analysing genomic data at scale, using the function *hl.import_vcf.* Subsequence sample- and variant-level quality control (QC) was performed using the Hail framework (see below), following mainly the workflows proposed by the gnomAD project[1], otherwise explicitly specified. The Hail-based pipelines used in this study are publicly available on GitHub (https://github.com/enriquea/wes_chd_ukbb).

### Sample QC

#### Hard filters.

To compute sample QC metrics, a set of high-confidence variants was defined by applying the following criteria: (i) bi-allelic, (ii) variants with high call-rate (> 0.99) across all samples in the call set and (iii) common single nucleonic variants (allelic frequency > 0.1%). The individual's chromosomal sex was inferred by calculating the inbreeding coefficient (*F-stat*) on chromosome X over the set of variants described above. The *hl.impute_sex* Hail function was used to perform the computation. This approach adopts the same implementation as the PLINK tool (v1.7). In addition, the coverage of the chromosome Y (normalized to chromosome 20) was used with the *F* stat to define the sample sex as follow: *male*: *F* > 0.6 and normalized Y coverage > 0.1, *female*: *F* < 0.4 and normalized Y coverage < 0.1. Samples with values outside these ranges were labelled as sex unspecific (**Supplemental data, Figure S1**). Samples were marked as failing hard filters if: a) chromosomal sex was unspecific, b) exhibited sample-specific low call rates (< 0.85) and c) mean coverage on chromosome 20 was equal to zero. **Table S1** (**Supplemental data**) summarises the number of samples affected per hard filter.

#### Inferring population ancestry.

The 1000 Genomes Phase 3 sequence data aligned to the human genome build GRCh38 (European Variation Archive (EVA) accession: PRJEB30460) was used to impute the global ancestry within the samples in the exome sequencing cohort. Both datasets were first merged based on locus and reference/alternate alleles. After merging, the Hail function *hl.hwe_normalized_pca* was used to compute the top 15 principal components on the subset of the well-behaved variants, defined as described above (see Hard filters section). A total of ~76,000 variants were included in the final set.

The set of 2,548 samples with known ancestry (from the 1000 Genomes Phase 3 dataset) was leveraged to build a random forest-based classifier using the top 15 computed principal components (PCs) as input features. Two-thirds of these samples were used as a *training dataset* and the remainder used as a *test dataset*. This step was combined with a recursive feature (a.k.a principal components) elimination procedure to define the optimal combination of PCs achieving the highest accuracy in the classification on the test data. In addition, a 10-fold cross-validation step was used for tuning the model parameters as previously described[2].

The model achieving the highest accuracy (>0.97) was then used to predict the ancestry of the remaining samples (discovery dataset with unknown ancestry). Each sample was broadly assigned to one of European (EUR), American (AMR), African (AFR), East Asian (EAS) or South Asian (SAS) population labels if random forest probability ($p$) > 0.8. Samples failing this threshold were labelled as OTHER. **Figure S2** and **Table S2** (**Supplemental data**) summarise the ancestry inference process results. The implemented approach showed high accuracy in classifying samples with reported ethnicity from the UK Biobank cohort (**Supplemental data, Table S3**).

#### Inferring sample relatedness.

The *hl.pc_relate* function from Hail was used to compute the relatedness between samples. Relatedness was computed among samples passing the hard filters. A variant was considered for inferring relatedness if it met the following criteria: 1) protein-coding exonic variant, 2) autosomal, 3) bi-allelic single nucleotide variants

(SNVs), 4) call rate across samples > 95%, 5) allele frequency (internal) > 1% and 6) LD-pruned with a cut-off at *r2* = 0.1. After running *hl.pc_relate*, Hail's *hl.maximal_independent_set* function was used to select the largest set of samples with no pair of samples related at the second-degree relatedness or closer (kinship coefficient > 0.125), prioritising cases over controls. This process filtered out a total of 3,782 samples (either twin/duplicated or first-degree relatives).

#### Platform inference.

Detailed capture platform meta-data information was missing for a fraction of the samples within the assembled cohort (~20%). To impute a platform for these samples, we adopted the data-driven approach proposed by gnomAD[1]. In brief, a list of the known exome capture intervals across multiple exome capture products was compiled for imputing samples platforms (including Agilent Sure Select All Exons products (version 2 to 5) and IDT xGEN). Only bi-allelic variants falling within these regions were included in the analysis. A sample per interval call-rate matrix was computed by considering the set of biallelic variants within each interval. The call-rate values were further discretised as non-called (0) and called (1) by applying a call-rate cut-off at 0.25 and principal component analysis performed on the discrete matrix. The top seven principal components (variance explained higher than 98%) were used as input for HDBSCAN (https://hdbscan.readthedocs.io), an unsupervised clustering method that allowed us to group and assign generic sample platform labels. **Figure S3** shows the samples projected onto principal components two and three. This method assigned the platform accurately for 100% of the samples in the UK Biobank (those with known platform labels), demonstrating the validity of this approach.

### Platform- and population-specific outliers filtering.

Sample ancestry and capture platform are two of the most frequent cofounders when analysing exome sequencing data. Thus, we computed a set of sample quality control metrics stratified by population and platform to detect sample outliers. Specifically, we computed the number of deletions, the number of insertions, the number of SNVs, the ratio of deletions to insertions, the ratio of transitions to transversions, and the ratio of heterozygous to homozygous variants using the Hail function *hl.sample_qc*. A sample was marked as an outlier and filtered out if the value for a given QC metric was four median absolute deviations (MAD) from its median. **Table S4 (Supplemental data)** summarises the number of samples detected as outliers per evaluated QC metric.

### Final sample QC and evaluation.

After applying the above sample QC steps and filtering out the samples without approval for analysis, our cohort consisted of 49,308 samples (**Supplemental data, Table S5**). At this stage, multi-allelic variants were split using the Hail function *hl.split_multi_hts,* and the dataset was filtered to high-quality genotypes. Genotypes were defined as high-quality if: a) dept of coverage >= 10, b) genotype quality >= 20 and c) genotype allele balance of heterozygotes > 0.20.

In addition, we evaluated the per sample distribution of the depth of coverage (DP) and genotype quality (GQ) stratified by case/control and male/female status. Our analysis revealed a comparable distribution of these metrics between cases/controls (**Supplemental data, Figure S4**) and male/females (**Supplemental data, Figure S5**). Mean DP values ranged between 20-35X (recommended cut-off is >10X) whereas GQ values ranged between 50-80 (recommended cut-off is >20).

### Variant QC

To define a set of high-quality variants for downstream analysis, we then applied several QC steps to the variants present in samples passing the sample QC process.

#### Hard filters.

We followed the variant QC scheme proposed by Karczewski *et al.*[1], where variants were flagged as failing hard filters if they showed a) an excess of heterozygotes (inbreeding coefficient < -0.3) and b) an absence of at least one sample with a high-quality genotype (allele-count zero, as defined above).

#### RF model.

A random forest (RF) model was trained and applied to distinguish true variations from potential false positives[1]. Positive training sets were downloaded from gnomAD repository (gs://gcp-public-data--gnomad/truth-sets/hail-0.2). Variants failing traditional GATK hard filters (QD < 2 or FS > 60 or MQ < 30) were used as a negative training set. Allele- and site-specific sequencing quality metrics were used as features for training the model (**Supplemental data, Table S6**). Features were imputed using its median where the value was missing. The chromosome 20 (test set) was left out of the training process for evaluation proposes. The final RF model achieved an accuracy >0.97 on this set of variants (test set). A variant was filtered out if the RF probability of being false positive was higher than 0.8.

#### VQSR filter.

In addition to the proposed RF model, we applied the conventional GATK Variant Quality Score Recalibration (VQSR) as a complementary approach to filter out low-quality variants. We used the recommended annotations and training datasets as suggested by the GATK best practices (https://gatkforums.broadinstitute.org/gatk). Both SNVs and indels were excluded if they failed the VQSR filter, according to the default settings. This allowed us to identify a fraction of variants that were likely false positives that passed the RF filter (**Supplemental data, Figure S6**).

#### Coverage.

Finally, we defined a variant as passing the QC if a) was covered by the major capture platforms used in the assembled cohort (different versions of Agilent Sure Select All Exome and IDT xGen panel 1 and b) showed coverage of 10X or more in at least the 90% of the samples in the gnomAD genome dataset (version 3.1.0).

**Table S7 (Supplemental data)** summarises the number of variants affected by each applied filter and the final number of variants considered for further analysis.

### Variant annotation

The cohort-VCF file was annotated using the Variant Effect Predictor tool (API version 94) with the flag *--everything*. The most severe variant consequence per protein-coding transcript was considered. The variant consequence severity was set based on the severity rank from Ensembl (https://www.ensembl.org), which prioritise variants as follows: protein-truncating > protein-altering > synonymous variants. The VEP tool functionalities were extended by using the plug-ins CADD (version 1.6) and dbNSFP[3] (version 4.1a) to annotate different missense variant pathogenicity scores (CADD[4], MPC[5], REVEL[6] and MVP[7]).

### Supplemental Data (Sample and Variant QC)

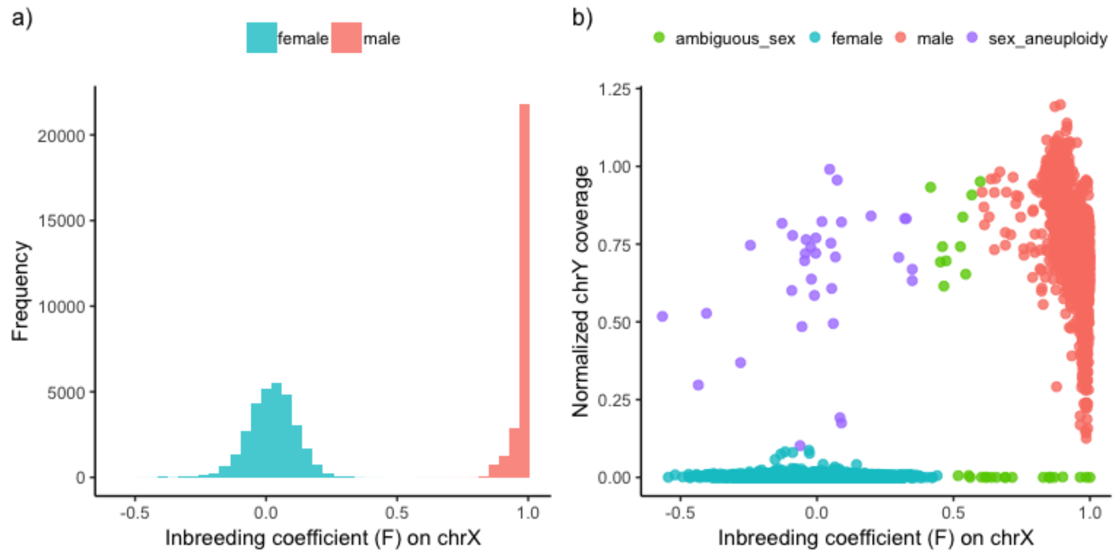

**Figure S1.** a) Inbreeding coefficient (F-stat) distribution computed over 57,628 samples. b) Inbreeding coefficient (x-axis) vs. normalized chromosome Y coverage (y-axis). Sample chromosomal sex was defined as follow, i) female: F < 0.4 and coverage chrY < 0.1, ii) male: F > 0.6 and coverage chrY > 0.1, iii) aneuploidy: F < 0.4 and coverage chrY >= 0.1, iv) samples failing any of these criteria were flagged as 'ambiguous sex'.

**Table S1**. The number of affected samples per hard filter.

| **Hard filters** | **N. of samples** | **Percent (%)** |
|---|---|---|
| Low call rate | 9 | 0.02 |
| Low coverage | 1 | 0.00 |
| Ambiguous sex | 30 | 0.05 |
| Sex aneuploidy | 34 | 0.06 |
| Filters combined | 72 | 0.12 |

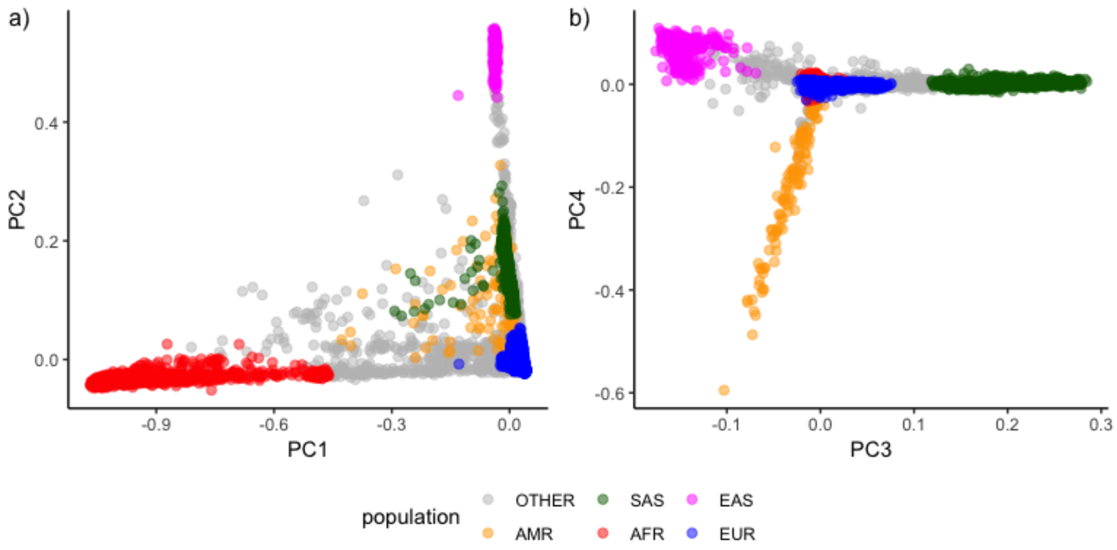

**Figure S2.** Samples projected onto the top four ancestry principal components (PCs) and their classification into five major ancestral populations. Samples were assigned to SAS, EAS, AMR, AFR or EUR if random forest probability (*p*) > 0.8. Samples failing this threshold were labelled as OTHER (grey). a) PC1 vs PC2 and b) PC3 vs PC4.

**Table S2.** The number of samples assigned per population. As expected, most samples were assigned to European ancestries (~91%). Approximately 3% of the samples were not assigned to a specific population (labelled as OTHER).

| Population | N. of samples | Percent (%) |
|---|---|---|
| AFR | 1,196 | 2.07 |
| AMR | 111 | 0.19 |
| EAS | 313 | 0.54 |
| EUR | 52,844 | 91.63 |
| OTHER | 1,772 | 3.07 |
| SAS | 1,437 | 2.49 |

**Table S3**. Confusion matrix with assigned population vs reported ethnicity for samples from the UK Biobank (UKBB).

| Assigned population | Reported ethnicity | N. of samples per assigned population | N. of samples per reported ethnicity | Percent true classified (%) |
|---|---|---|---|---|
| AFR | African | 319 | 332 | 96.08 |
| EUR | British | 42450 | 43184 | 98.30 |
| EAS | Chinese | 169 | 173 | 97.69 |
| SAS | Indian | 690 | 708 | 97.46 |
| EUR | Irish | 1495 | 1498 | 99.80 |
| SAS | Pakistani | 138 | 138 | 100.00 |

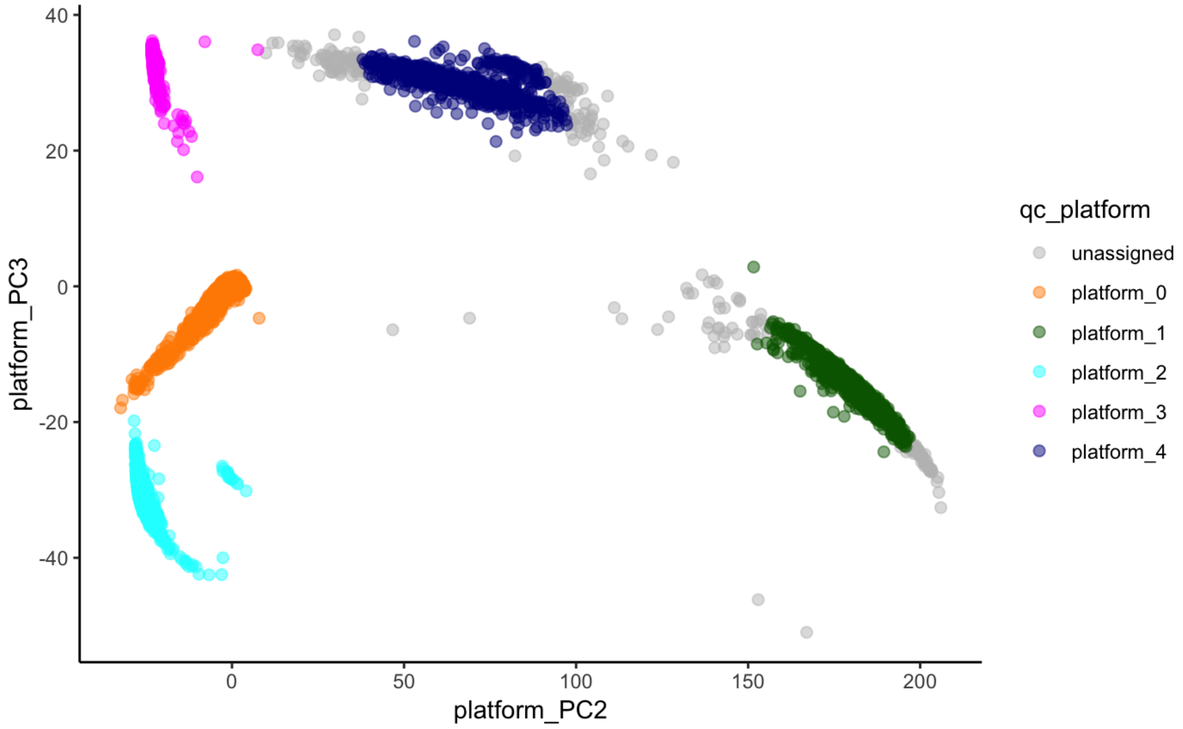

**Figure S3.** Samples projected onto the platform's principal components (PC) two and three. No generic platform (grey dots) was assigned for less than 0.5% of the samples (n=233). The proposed clustering approach accurately assigned 100% of the samples in the UK Biobank cohort (samples with known capture platform information, orange cluster). The exome capture platform intervals used in the analysis are described in the Methods section.

**Table S4.** The number of samples detected as outliers by evaluating different sample quality control (QC) metrics. Samples were grouped as per assigned population/platform, and QC metrics were computed per group. Multiple samples (n=104) were detected as outliers by two or more QC metrics.

| QC metrics | N. of sample outliers | Percent (%) |
|---|---|---|
| Number of SNPs | 134 | 0.23 |
| Number of deletions | 85 | 0.15 |
| Number of insertions | 85 | 0.15 |
| Ratio transmission/transversion | 89 | 0.15 |
| Ratio insertion/deletion | 14 | 0.02 |
| Ratio heterozygous/homozygous | 266 | 0.46 |

**Table S5.** Number of remaining samples after each filter stage. *Population filter refers here to samples with assigned European ancestries.

| Filter stages | Remaining samples |
|---|---|
| Unfiltered | 57,628 |
| Hard filters | 57,560 |
| Hard filters, relatedness | 53,862 |
| Hard filters, relatedness, QC outliers | 53,507 |
| Hard filters, relatedness, QC outliers, *population | 49,308 |

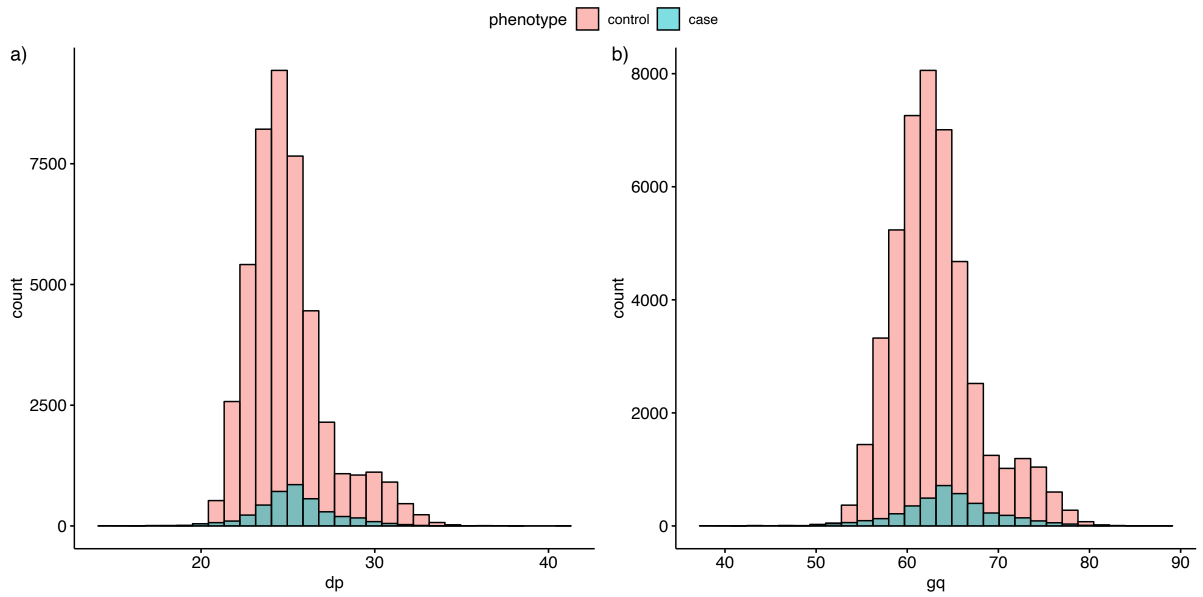

**Figure S4.** Distribution of per sample averaged QC metrics stratified by phenotype (case/control). a) Mean depth of coverage (DP) and b) Mean genotype quality (GQ). QC metrics were computed per sample across autosomal variants.

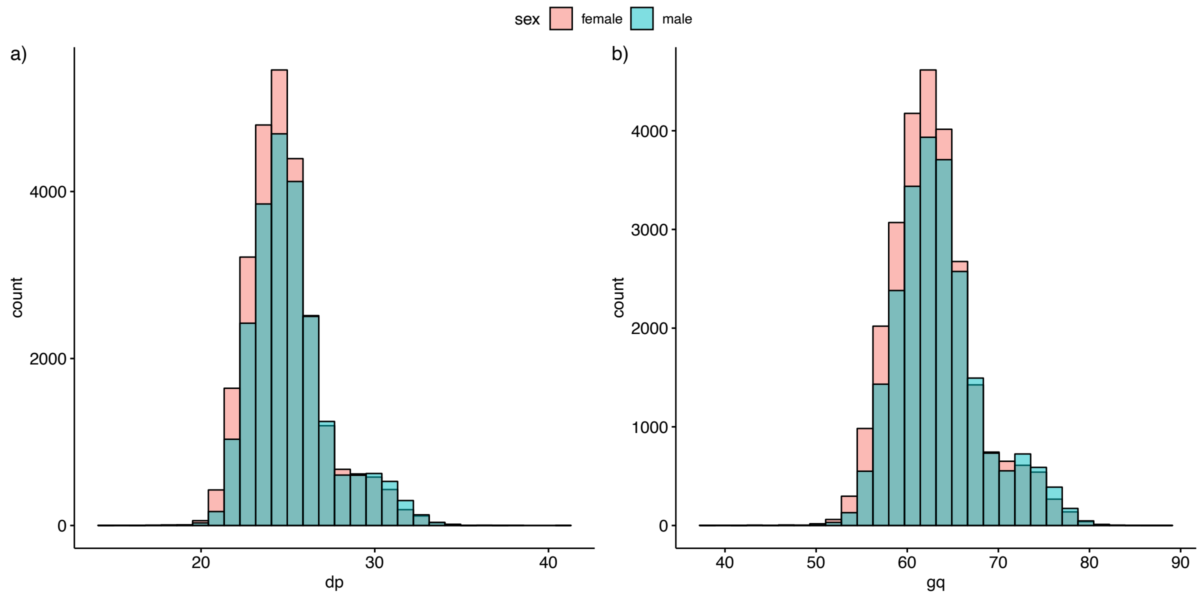

**Figure S5.** Distribution of per sample averaged QC metrics stratified by sex (female/male). a) Mean depth of coverage (DP) and b) Mean genotype quality (GQ). QC metrics were computed per sample across autosomal variants.

**Table S6.** Features used in the random forest model to predict the variant probability of being true positive or false positive.

| RF features | Description | Importance |
|---|---|---|
| variant_type | SNV or indel | 0.011 |
| SOR | Symmetric Odds Ratio of 2x2 contingency table to detect strand bias | 0.105 |
| ReadPosRankSum | Z-score from Wilcoxon rank-sum test of Alt vs Ref read position bias | 0.016 |
| InbreedingCoeff | Inbreeding coefficient as estimated from the genotype likelihoods per-sample when compared against the Hardy-Weinberg expectation | 0.103 |
| FS | Phred-scaled p-value using Fisher's exact test to detect strand bias | 0.041 |
| DP | Approximate read depth | 0.003 |
| QD | Allele-specific Variant Confidence/Quality by Depth | 0.704 |
| was_mixed | True if both SNVs and indels are present at the site | 0.001 |
| n_alt_alleles | Number of alleles at the site | 0.001 |
| MQRankSum | Z-score From Wilcoxon rank-sum test of Alt vs Ref read mapping qualities | 0.010 |

**Table S7.** The number of remaining variants per filter stage. RF: Random Forest filter, VQSR: Variant Quality Score Recalibration, Coverage: >10X in at least 90% of the samples in gnomAD genome cohort.

| **Filter stages** | **Remaining variants** |
|---|---|
| Unfiltered | 11,433,645 |
| Hard filters | 11,406,658 |
| Hard filters, RF | 9,490,151 |
| Hard filters, RF, VQSR | 9,191,448 |
| Hard filters, RF, VQSR, Coverage | 9,134,464 |

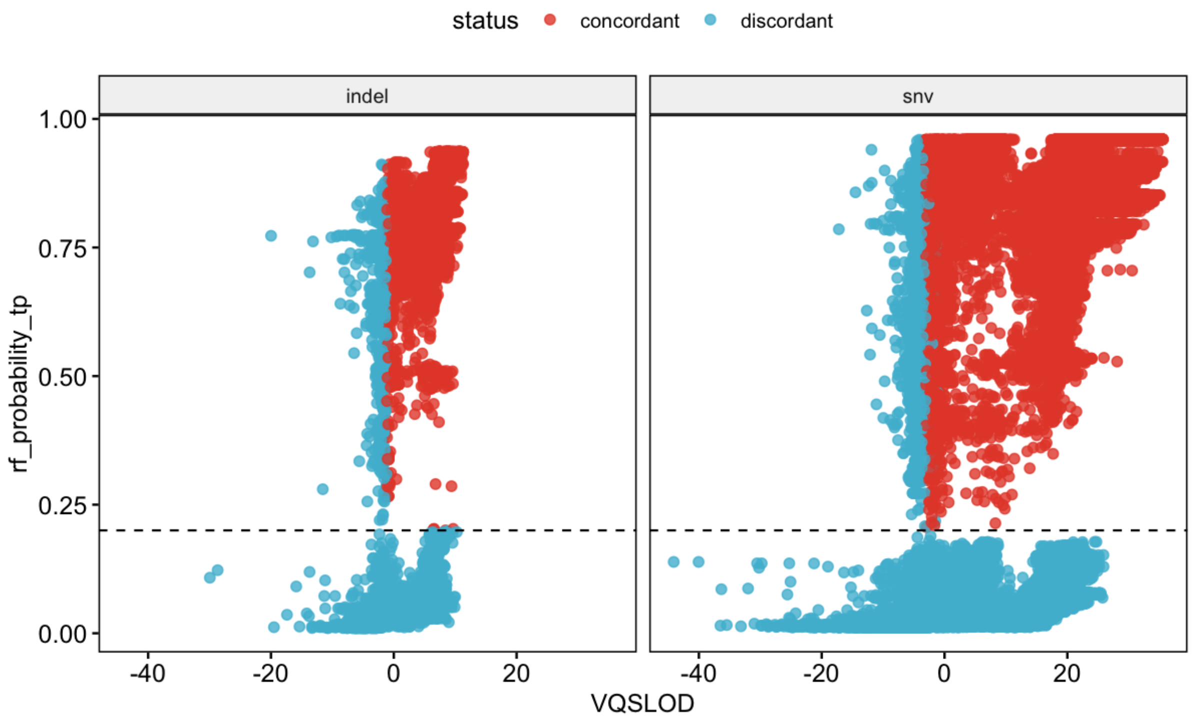

**Figure S6.** Variant quality score recalibration (x-axis) vs Radom Forest (RF) probability of being true positive (y-axis). Variants (SNVs and indels) are depicted for chromosome 20. The dashed line indicates the cut-off used for the RF probability (=0.2). Concordant (red dots): variants pass both the RF and VQSR filters; discordant (blue dots): variants fail at least one of the RF or VQSR filters.
